# Reduced brain structural similarity is associated with maturation, neurobiological features, and clinical status in schizophrenia

**DOI:** 10.1101/2024.11.25.24317878

**Authors:** Natalia García-San-Martín, Richard AI Bethlehem, Patricia Segura, Agoston Mihalik, Jakob Seidlitz, Isaac Sebenius, Claudio Alemán-Morillo, Lena Dorfschmidt, Golia Shafiei, Sarah E. Morgan, Miguel Ruiz-Veguilla, Rosa Ayesa-Arriola, Javier Vázquez-Bourgon, Bratislav Misic, John Suckling, Benedicto Crespo-Facorro, Rafael Romero-García

**Author notes:** Correspondence to: Rafael Romero García, Full address: Avda. Doctor Fedriani S/N, 419009, Seville (Spain).

## Abstract

Schizophrenia spectrum disorders (SSD) are characterized by atypical brain maturation, including alterations in structural similarity between regions. Using structural MRI data from 195 healthy controls (HC) and 352 individuals with SSD, we construct individual Morphometric INverse Divergence (MIND) networks. Compared to HC, individuals with SSD mainly exhibit reduced structural similarity in the temporal, cingulate, and insular lobes, being more pronounced in individuals exhibiting a ‘poor’ clinical status (more impaired cognitive functioning and more severe symptomatology). These alterations are associated with cortical hierarchy and maturational events, locating MIND reductions in higher-order association areas that mature later. Finally, we map 46 neurobiological features onto MIND networks, revealing a high presence of neurotransmitters and astrocytes, along with decreased metabolism and microstructure, in regions with reduced similarity in SSD. These findings provide evidence on the complex interplay between structural similarity, maturational events, and the underlying neurobiology in determining clinical status of individuals with SSD.

## Introduction

Multiple MRI studies have detected brain structural alterations across several cortical areas in individuals with schizophrenia spectrum disorders (SSD), including reductions in volume, surface, thickness, and gyrification^1–5^. The widespread, multicentric pattern of these structural alterations likely reflects dysconnectivity within brain specific networks^6^. The structural connectome, traditionally derived from diffusion-weighted imaging (DWI), has provided valuable insights into connectivity alterations associated with SSD^7,8^. For example, a more efficiently wired connectome has demonstrated a better response to antipsychotic treatment^8^. However, this tractography technique generates a substantial amount of false-positive fiber bundles^9^. Alternatively, structural networks can be constructed using within-subject cortical similarity approaches, which have shown a correspondence with tract-tracing similar to that observed in previous reports on DWI-based networks in macaques^10,11^. Additionally, similarity networks have demonstrated potential for clinical translation due to their reduced acquisition times, as networks derived from a single, routinely acquired T_1_-weighted MRI scan have been shown to correspond to those generated from a broader set of multimodal MRI features^12^. These networks have also demonstrated to be sensitive to the duration of psychosis^13^, the expression of schizophrenia-related genes^6^, and the prediction of treatment response^14^, providing novel insights into the role of brain structure in mediating genetic and neuronal factors that underlie clinical outcomes and the progression of the disease. Moreover, a recent method based on Morphometric Inverse Divergence (MIND) has emerged, providing an estimate of within-subject similarity between cortical areas based on multiple MRI features^15^. These novel similarity networks have demonstrated increased inter-individual consistency, stronger inter-hemispheric and within-class connections, and increased coupling with gene co-expression compared to DWI tractography^15^. Furthermore, MIND networks have also shown greater reliability, better consistency with cortical cytoarchitectonics and symmetry, and stronger correlations with tract-tracing measures of axonal connectivity compared to previous approaches^15^. Exploring MIND networks among individuals with SSD may shed light on the complex structural alterations that manifest as diverse cognitive and symptomatologic status.

Extensive research has linked volumetric reductions and structural network alterations to severe symptomatology in first-episode psychosis (FEP) and chronic schizophrenia^5,16,17^. Investigations on drug-naïve patients are scarcer, but they have reported decreases in cortical thickness and structural connectivity associated with cognitive and functioning deficits^18–20^. In this context, baseline reductions of structural connectivity have successfully predicted worse functioning outcomes in antipsychotic-naïve FEP individuals, which could not be attributed to antipsychotic medication^19^. Additionally, reduced overall DWI connectivity and longer path length have also predicted lower general cognitive ability in individuals with schizophrenia^21^. Positive symptoms have also been associated with a widespread pattern of decreased gray matter volume in bilateral frontotemporal and occipital regions in FEP individuals^5^. On the other hand, structural alterations associated with negative symptomatology have shown inconsistencies^22^. The controversial impact of antipsychotic drugs on symptomatology and brain architecture^23–25^ highlights the relevance of research on drug-naïve or minimally medicated patients, which could help elucidate the specific contribution of disease-related alterations to brain structure.

A growing body of literature supports the hypothesis that psychiatric and neurological conditions initially manifest as abnormal brain structures in specific regions, referred to as epicenters^26,27^. These abnormalities subsequently spread to other areas through the underlying network architecture, involving genetic, trans-neuronal, structural, and functional processes^16,28–30^. These mechanisms may disrupt the transport of trophic factors and facilitate the spread of misfolded proteins or inflammatory markers linked to cell death and atrophy, contributing to the structural and functional progression of multiple neurodegenerative diseases^31–33^. Moreover, epicenters have been found to correlate with more severe symptom profiles, suggesting that network organization may guide cortical alterations throughout the disease process of schizophrenia and may be influenced by cognitive impairment and symptom severity, among other factors^16^. However, the pathological spread of these abnormalities from disease epicenters to connected brain regions, resulting in network-wide morphological changes, remains unclear. Investigating the relationship between network architecture and atypical development in SSD may be key to understanding abnormal brain maturation and functioning^34^.

A key aspect of human brain maturation is the consolidation of cortical hierarchy, where higher-order association areas exhibit faster rates of shrinkage and myelination compared to low-order sensorimotor areas during the course of adolescence, facilitating efficient connectivity throughout the structural network^35^. Population models have become a fundamental tool for accurately delineating these brain maturational events, including determining the ages at which regions reach their peak volume and maximum growth velocity^36^. These approaches based on normative data also enable benchmarking regional brain volumes to identify patterns of atypical neuroanatomical maturation after accounting for age-, sex-, and site-effects^36^. This allows for constructing population-relative ‘centiles’, a measure of brain structure atypicality across the lifespan. A recent study based on this methodology demonstrated common abnormal trajectories of cortical volume across different psychosis-related conditions, supporting the notion that regions with similar structural profiles are co-vulnerable to psychosis (i.e., exhibit similar psychosis-related centile alterations)^37^. Although advanced structural brain ageing^38^ and alterations in higher-order areas^39,40^ have been implicated in schizophrenia, no study has yet explored the complex interaction between maturational events, brain hierarchy, and structural connectivity derived from similarity networks in SSD.

Understanding how structural alterations in psychiatric conditions relate to the underlying neurobiological substrates has been particularly challenging due to limited human data^41,42^. However, recent studies have explored the spatial distribution of several neurobiological properties, including molecular and cellular markers^43,44^. One study revealed that, while psychotic-related conditions exhibit distinct patterns of gray matter reductions, there is a major overlap in the neurobiological features characterizing atypically maturing regions, including a higher density of serotonin receptors, among others^37^. No study to date has explored the biological profiles associated with structural alterations in network connectivity and their potential implication for SSD clinical status^6^.

Here, we exploit a method based on MIND networks to assess how structural markers are associated with abnormal development in SSD and with their clinical status. We constructed MIND networks by estimating within-subject similarities between cortical areas in both SSD and healthy control (HC) subjects. Firstly, we compared the differences in MIND similarity (also referred here as connectivity) between SSD and HC, and then after stratifying individuals with SSD into ‘poor’ or ‘good’ clinical status. Secondly, we examined whether any decrease in MIND connectivity of patients with SSD was linked to centile reductions or other maturational events. For this purpose, we first computed the regional connectivity to disease epicenters of these individuals to assess how clinical status was influenced by the regions preferentially linked to atypically maturing areas. Next, we explored potential associations between MIND networks and the hierarchical organization of the cortex. Finally, we conducted a multivariate analysis to examine the associations between regional neurobiological features and MIND alterations in relation to clinical status.

## Results

### Decreased MIND connectivity in schizophrenia is associated with clinical status

MIND networks were computed as the within-subject similarity between cortical areas based on the Kullback-Leibler divergence of multiple MRI features (see *Methods* for details). To determine how MIND varies between SSD and HC, we computed the effect size (Cohen’s d) between networks (for edges and nodes separately). Compared to HC, individuals with SSD exhibited significant reductions in MIND connectivity in edges targeting the temporal, cingulate, and insular cortices (*P*_perm_ < 0.05), while edges interconnecting the occipital cortices showed higher connectivity (Fig. 1A, left and right; see Source Data for the full matrix). Frontal and parietal connecting regions exhibited both increases and decreases in MIND connectivity. At the regional level, patients with SSD exhibited a significant reduction in MIND degree in the insular, cingulate, and temporal cortices, along with an increase in the occipital lobe of the right hemisphere (Fig. 1A middle and right). The sensitivity analyses revealed that MIND degree remained highly consistent across both scanning sites and sexes (see Supplementary Fig. 1), and was robust to inter-individual variations, image quality (Euler index), and parcellation schemes (Supplementary Fig. 2A-C). Effect sizes were also significantly correlated with those reported by Morgan *et al*.^6^ in Morphometric Similarity (MS) networks derived from individuals with chronic schizophrenia (*r*_partial_ = 0.35, *P*_spin_ = 0.004; Supplementary Fig. 2D).

**Figure 1.**
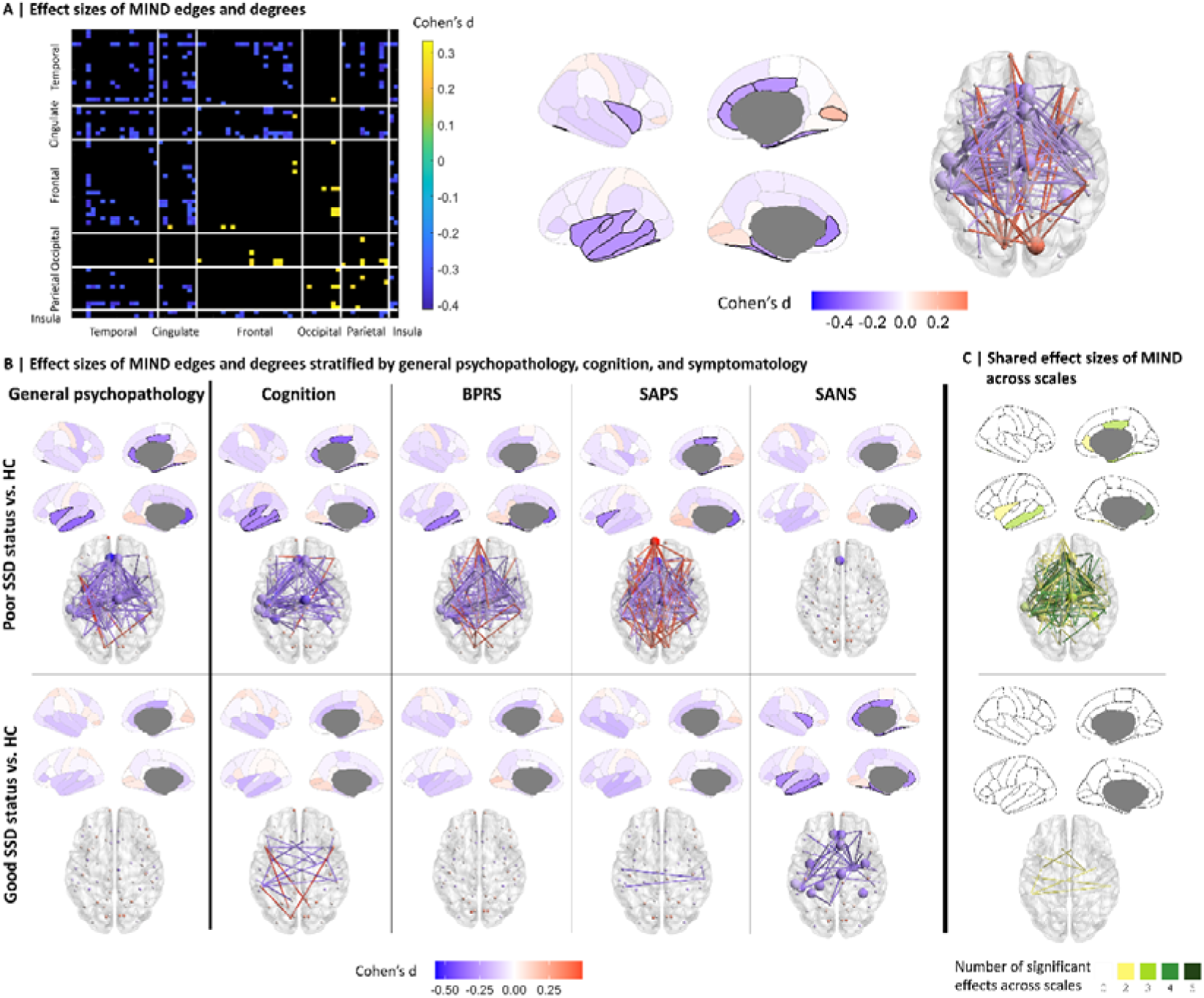
MIND connectivity in relation to clinical status. (**A**) Effect sizes (Cohen’s d) computed between SSD and HC MIND edges (left) and degrees (middle), where blue indicates a significant reduction of edge and degree in SSD, yellow and red indicate a significant increase, and black indicates no significant differences. Spatial 3D representation of significantly different edges and degrees in SSD compared to HC (right). Supplemental data are provided as a Source Data file. (**B**) Effect sizes of MIND edges and degrees stratified by general psychopathology, cognition, and symptomatology. The Cohen’s d of MIND degree for those individuals with SSD whose general psychopathology is more impaired, whose cognition is lower, and whose symptomatology (BPRS, SAPS, and SANS) is equal or higher than the median of the sample (top row). The Cohen’s d for those whose general psychopathology is less impaired, whose symptomatology is lower, and whose cognition is equal or higher than the median of the sample is shown in the bottom row. The highlighted regions show those regional degrees that exhibit significant differences from HC after tail area-based false discovery rate (FDR) correction (permutation test, *P*LJ<LJ0.05). (**C**) Number of significant effect sizes shared across scales for a ‘poor’ and ‘good’ status. BPRS, Brief Psychiatric Rating Scale; HC, healthy control; MIND, Morphometric INverse Divergence; SANS, Scale for the Assessment of Negative Symptoms; SAPS, Scale for the Assessment of Positive Symptoms; SSD, schizophrenia spectrum disorder.

Effect sizes of MIND edges and degrees were analyzed based on the general psychopathology and individual scores of global cognitive functioning and clinical symptoms (BPRS, SAPS, and SANS scales) of patients with SSD, which were classified with a ‘poor’ or ‘good’ status (see *Methods* and Supplementary Fig. 3 for details on the score distributions across each status; see Supplementary Fig. 4 for regional correlations between MIND degree and clinical status). Instead of reporting individual scale scores for cognition, we used an average score for simplicity. This decision was supported by the Principal Component Analysis (PCA) performed on these scales, where the first principal component explained 58.65% of the variance and showed highly similar weights across scales (ranging from 0.27 to 0.44). Compared to HC, alterations in MIND were observed only in patients with more impaired general psychopathology (‘poor’ general psychopathology; Fig. 1B). Consistent with these findings, reductions in MIND degrees (cingulate, fusiform, and temporal regions) were observed only in patients with more impaired cognitive functioning (‘poor’ cognition). Accordingly, patients with more severe symptomatology (‘poor’ clinical status) exhibited a significant reduction in MIND degree in the left rostral anterior cingulate region across all three scales (Cohen’s d < −0.32), but not in individuals with less severe clinical symptomatology (‘good’ clinical status), except for SANS. Additionally, individuals with higher BPRS total scores showed several significant reductions in MIND edges connecting frontal, temporal, and cingulate regions (Cohen’s d < −0.28), along with few increases in edges connecting frontal and occipital regions (Cohen’s d > 0.28). However, the lowest score individuals did not show any significant difference compared to HC. While individuals with higher SAPS scores presented the temporal and cingulate edges significantly reduced (Cohen’s d < −0.27), as well as the frontal and occipital edges increased (Cohen’s d > 0.27), individuals with lower SAPS scores barely presented differences compared to HC. Conversely, individuals with lower SANS scores presented a greater number of significantly reduced regions (temporal, insular, and cingulate cortices) compared to individuals with higher scores. Similar results were obtained when Desikan-Killiany-318 atlas parcellation was employed (Supplementary Fig. 5). The number of significant effect sizes shared across scales was higher for individuals with ‘poor’ status (Fig. 1C; see Supplementary Fig. 6 for the distinct effect sizes of MIND edges and degrees of each scale for a ‘poor’ and ‘good’ status).

Additionally, the association between MIND degree and clinical status was also explored without stratifying individuals into ‘poor’ or ‘good’ status, supporting the finding that regions with the greatest reductions in MIND degree in SSD were those showing the strongest correlations with cognition (Supplementary Fig. 7).

### Connectivity to disease epicenters is associated with clinical status

In addition to the MIND network analyses, we identified the regions with the most pronounced morphological alterations at SSD onset (centiles). Regional centiles were computed for each participant by benchmarking regional MRI volumes against previously established normative trajectories^36^. These centiles were highly consistent across diagnostic groups (Supplementary Fig. 8). The SSD group exhibited a generalized significant centile reduction in almost all regions (Fig. 2A left), which were negatively correlated with HC degrees (*r* = −0.45; *P*_spin_ = 0.001). However, this association was no longer significant after accounting for regional volumes (Fig. 2A middle; *r*_partial_ = −0.20; *P*_spin_ = 0.122). To assess the relationship between MIND edges and the inter-regional susceptibility to psychosis, we computed the structural co-vulnerability matrix by correlating the regional effect sizes of the centiles for different psychosis-related groups (see *Methods* for details). Results revealed a significant association between MIND connectivity and inter-regional co-vulnerability, indicating that pairs of regions that were more structurally similar exhibited higher co-vulnerability to psychosis (Fig. 2A right; *r* = 0.33; *P*_Betzel_ < 10^-4^; *P*_Moran_ = 0.001; see Subjects and Methods for Betzel rewiring null model and Moran spectral randomization). Building on this edge-level association between MIND and centile-based alterations, we further analyzed the connectivity to disease epicenters.

**Figure 2.**
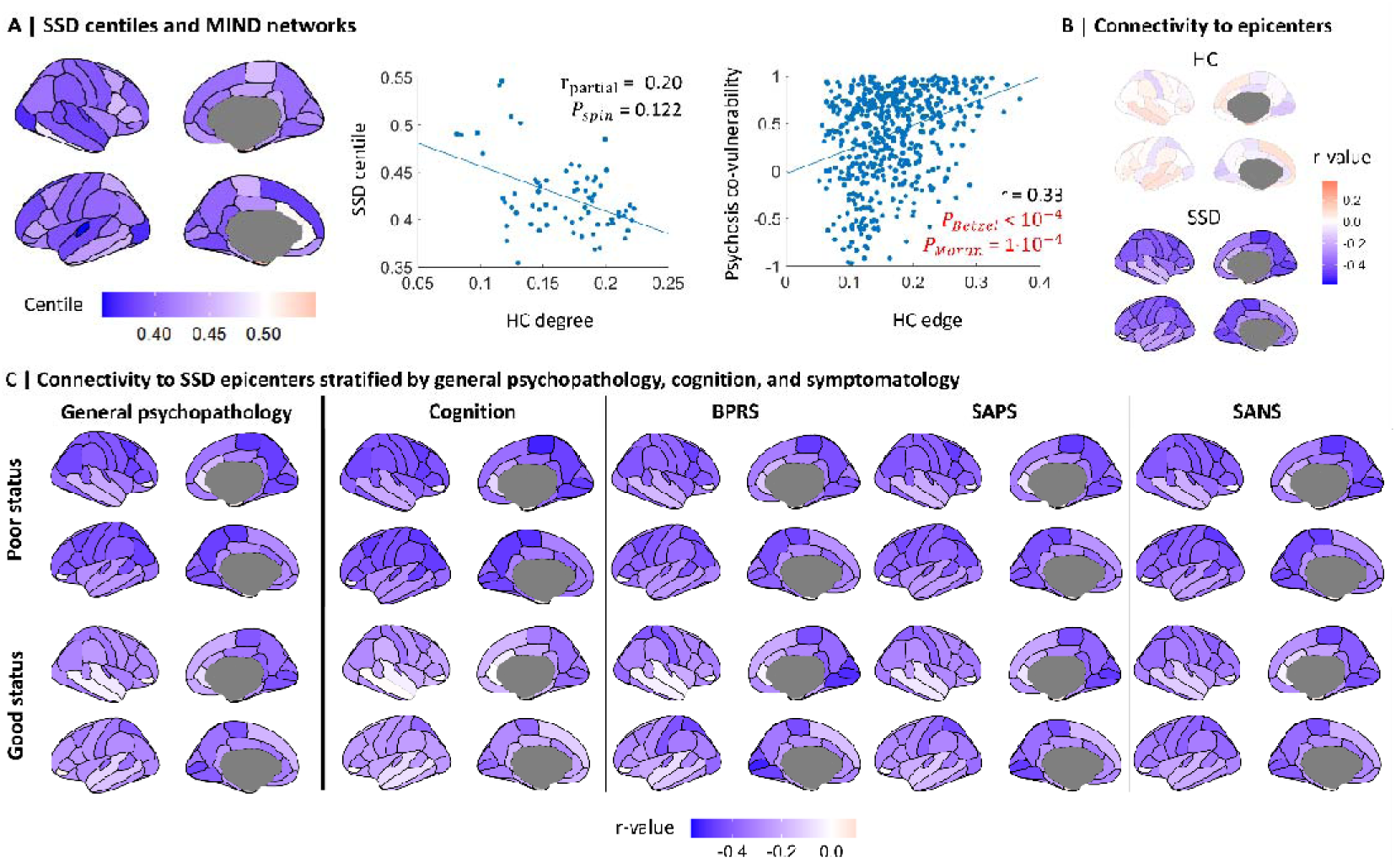
Connectivity to disease epicenters. (**A**) Regional MRI brain volumes from subjects with SSD converted into centiles, where blue and red indicate a significant centile reduction and increase, respectively, in SSD compared to the expected regional volume for age- and sex-matched normative individuals (left). Correlation between HC degrees and SSD (middle; two-sided test). Association between HC edges and the structural co-vulnerability to psychosis matrix (right; two-sided test). (**B**) Correlation between the corresponding edges to each region and the mean HC (top) and SSD centiles (bottom), where blue and red indicate a negative and positive correlation, respectively. (**C**) Connectivity to SSD epicenters stratified by patients with ‘poor’ and ‘good’ general psychopathology, cognition, and symptomatology (BPRS, SAPS, and SANS). The highlighted areas indicate regions where connectivity is significantly correlated with epicenters after tail area-based FDR correction (*P*_Betzel_lJ<lJ0.05; see Supplementary Fig. 12 for Moran significance). Supplemental data are provided as a Source Data file. BPRS, Brief Psychiatric Rating Scale; HC, healthy control; MIND, Morphometric INverse Divergence; SANS, Scale for the Assessment of Negative Symptoms; SAPS, Scale for the Assessment of Positive Symptoms; SSD, schizophrenia spectrum disorder.

Disease epicenters were defined as the set of regions with the most reduced centiles at SSD onset (see Supplementary Fig. 9 for correspondence with gray matter thickness abnormalities in chronic schizophrenia derived from ENIGMA^45^). According to Fig. 2A middle, these epicenters were significantly anti-correlated with the functional degree derived from an ENIGMA study in schizophrenia^16^ (*r* = −0.54, *P*_spin_ = 2·10^-4^, and *r* = 0.03, *P*_spin_ = 0.82, for functional and DWI connectivity, respectively; see Supplementary Fig. 10). To assess the structural similarity to these atypically maturating regions in SSD, the connectivity to epicenters was calculated for each region as the Pearson correlation between its edge (SSD or HC) and the corresponding SSD centiles (see *Subjects and Methods* for further details). Unlike HC epicenters, which did not reveal any significant association, several regions showed a significant negative correlation between edge strength and SSD centiles, indicating that strong edges are preferentially connected to SSD epicenters (Fig. 2B; *r_global_* = −0.27, computed as the average correlation across regions; see Source Data for correlation values along with their regional labels). After stratifying individuals by general psychopathology, individuals with ‘poor’ status exhibited a slightly stronger tendency to be connected to disease epicenters (*r_global_* = −0.29) compared to a ‘good’ condition (*r_global_* = −0.23; Fig. 2C). Accordingly, a moderately higher association between MIND and (*r_global_*= −0.33) disease epicenters was found in patients with ‘poor’ cognition compared to individuals with better cognitive functioning (*r_global_* = −0.16; see Supplementary Fig. 11 for regional correlations between disease epicenters and clinical status). Similarly, patients with more severe BPRS, SAPS, and SANS symptomatology exhibited the same tendency to be connected to epicenters (*r_global_* = −0.28, *r_global_* = −0.27, and *r_global_* = −0.28, respectively) compared to those with milder symptoms (*r_global_* = −0.21, *r_global_* = −0.22, and *r_global_* = - 0.24).

### Decreased MIND connectivity in schizophrenia is associated with cortical hierarchy and maturational events

We found no association between SSD centiles and the effect sizes of either MIND degree (Fig. 3A top; *r*_partial_ = −0.13; *P*_spin_ = 0.295) or edges (Supplementary Fig. 13; *r* = - 0.11; *P*_Moran_ = 0.43), revealing that atypical volumetric maturation and alterations in MIND network do not co-localize in the same cortical regions. Specifically, after correlating MIND degree with a sensorimotor-association regional ranking^46^, results showed that regions with reduced MIND degree in SSD were preferentially located in higher-order association areas (Fig. 3A middle; *r*_partial_ = −0.25; *P*_spin_ = 0.049), while SSD centiles were reduced in lower-order sensorimotor regions (Fig. 3A bottom; *r*_partial_ = 0.33; *P*_spin_ = 0.009). To determine whether MIND networks are related to maturational events, we exploited brain maps indicating the age at which maximum peaks of volume and velocity occur for each region^36^. Regions that reach maximum volume of maturation at an early age (e.g., frontal pole, superior parietal, and cuneus) exhibited stronger structural connectivity (hubs) in HC (Fig. 3B top; *r*_partial_ = −0.41; *P*_spin_ = 0.022). On the other hand, regions that mature later showed the strongest reduction in MIND degree in SSD compared to HC (Fig. 3B middle; *r*_partial_ = −0.53; *P*_spin_ = 0.002), but higher centiles in SSD (Fig. 3B bottom; *r*_partial_ = 0.63; *P*_spin_ = 0.001). Interestingly, although the peak age of regional velocity did not exhibit a significant correlation with HC degrees (Fig. 3C top; *r*_partial_ = 0.15; *P*_spin_ = 0.418), a significant association was found with the reduction in MIND degree of patients with SSD compared to HC (Fig. 3C middle; *r*_partial_ = −0.56; *P*_spin_ = 0.001), but not with SSD centiles (Fig. 3C bottom; *r*_partial_ = 0.10; *P*_spin_ = 0.602). All associations were calculated after accounting for mean regional volumes. Additionally, these findings remained highly consistent after stratifying individuals into ‘poor’ or ‘good’ status (Supplementary Figs. 14-17). Altogether, these findings reveal that reductions in regional structural similarity in individuals with SSD were co-localized with maturational events and higher-order association areas.

**Figure 3.**
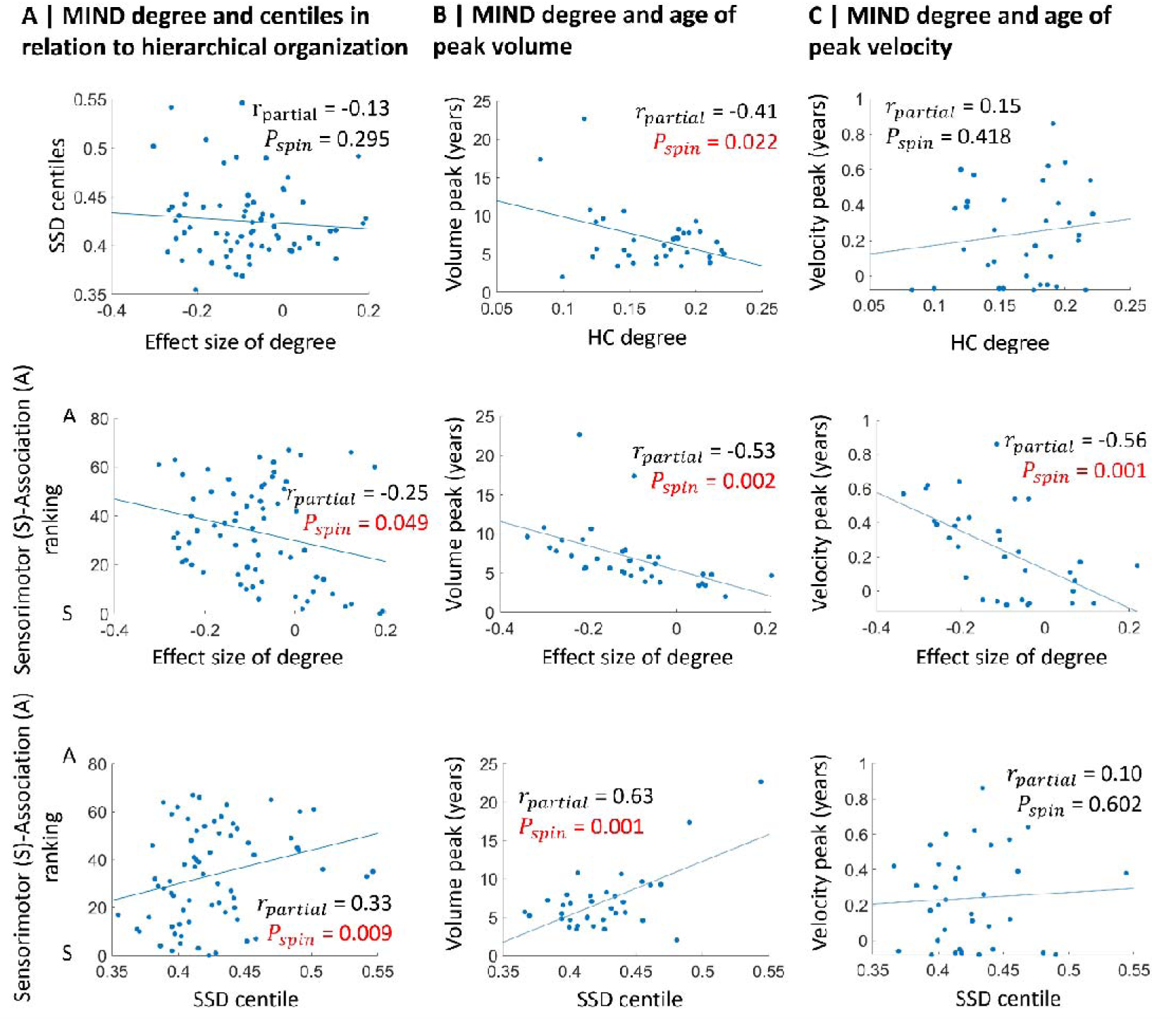
MIND connectivity in relation to sensorimotor-association hierarchy and maturational events. (**A**) Correlations between effect sizes of degrees and SSD centiles (top); effect sizes of degrees and cortical hierarchy (middle); SSD centiles and cortical hierarchy (bottom). (**B**) Correlations between the cortical map of age at regional volume peak (derived from normative trajectories) and HC degree (top); effect size of degree (middle); and SSD centiles (bottom). (**C**) Correlations between the normative cortical map of age at regional velocity peak and HC degree (top); effect size of degree (middle); and SSD centiles (bottom). All regional correlations (two-sided test) accounted for mean regional volumes. HC, healthy control; MIND, Morphometric INverse Divergence; SSD, schizophrenia spectrum disorder.

### Neurobiological features are co-localized with alterations in MIND connectivity and related to clinical status

To investigate the neurobiological features potentially linked to MIND networks, we exploited previously acquired datasets obtained from different modalities consisting of molecular and micro-architectural data clustered under different neurobiological types (neurotransmitter, cell type, layer thickness, microstructure, expansion, and metabolism; see *Subjects and Methods* for additional information)^47^. HC edges significantly correlated with the neurobiological similarity matrix, indicating that the more connected a pair of regions is, the more neurobiologically similar they are (Fig. 4A top; *r* = 0.47; *P*_Betzel_ < 10^-4^; *P*_Moran_ < 10^-4^). On the contrary, the effect size between SSD and HC edges did not correlate with neurobiology (Fig. 4A bottom; *r* = 0.11; *P*_Moran_ = 0.33). However, when this analysis was conducted separately for each neurobiological type, significant associations were identified with both HC and effect sizes (Supplementary Fig. 18). At the regional level, we performed a combined PCA and Canonical Correlation Analysis (CCA) approach to investigate the regional associations of MIND degree with the neurobiological features (see *Subjects and Methods* and Supplementary Fig. 19 for details). The PCA-CCA models were significant (FDR-corrected) for both HC (Fig. 4B top-left; r = 0.67; *P*_spin_ = 0.001) and effect sizes of degree (Fig. 4B bottom-left; r = 0.71; *P*_spin_ = 0.001). The predicted HC degrees and effect sizes resembled their corresponding empirical values (Fig. 4B middle). Both models exhibited significant loadings (Fig. 4B right; *P*_spin_ < 0.05). Cell types (Neuro-Ex/In), cortical expansion (Scaling PNC and NIH, developmental and evolutionary exp.), neurotransmitters (5-HT_2A_, 5-HT_4_), and metabolism (CMRGlu, Glycolytic index, CMRO_2_) demonstrated a significant positive contribution to HC degrees, indicating that the presence of these neurobiological features is highly co-localized with MIND hubs. The negative contribution of neurotransmitters (5-HTT, DAT, VAChT, 5-HT_1A_, NMDA, H_3_) and cell types (OPC, Micro, Astro) indicated a high presence of these features in less connected regions. The features that most strongly accounted for explaining the regional reductions in MIND degree between SSD and HC included neurotransmitters (D_2_, 5-HT_1A_, CB_1_, MOR, 5-HT_4_, DAT, 5-HTT, D_1_, mGluR_5_, NMDA, 5-HT_6_), cell types (OPC, Astro), and layer thickness (Layer I-III), indicating a high presence in regions with reduced SSD connectivity. Similar loadings were found when the cohort was randomly split into two halves (see Supplementary Fig. 20). On the other hand, a high presence of microstructural features (Gene PC1, Neurotransmitter PC1) and metabolism (CMRO_2_, CMRGlu, CBF) contributed to explaining the increased structural connectivity of individuals with SSD compared to HC. The neurobiological loadings that contributed to explaining the SSD centiles were shown in a previous study^37^.

**Figure 4.**
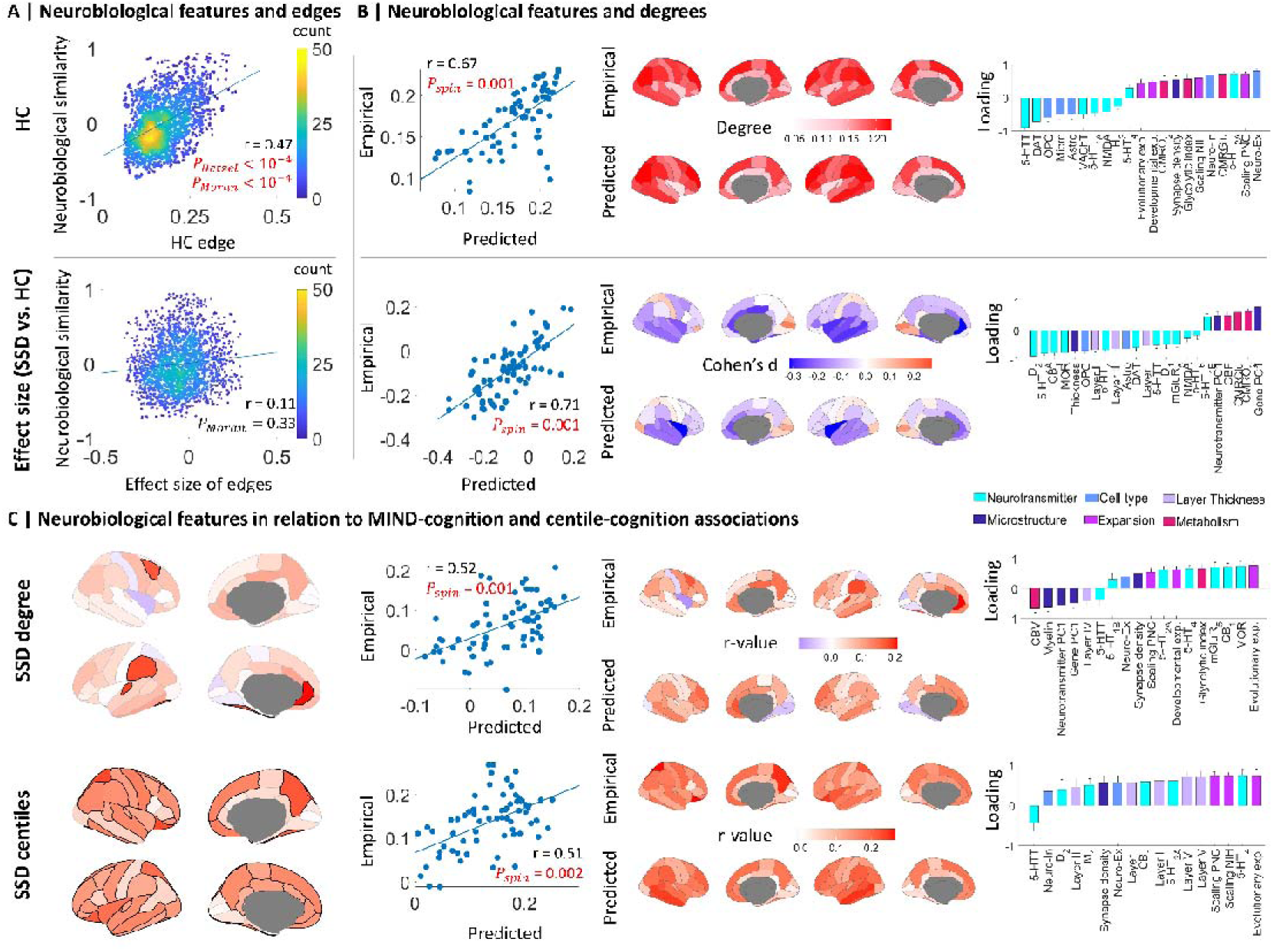
Neurobiological characterization of MIND networks. (**A**) Association (two-sided test) between the neurobiological similarity matrix and HC edges (top) and the effect sizes of MIND edges between SSD and HC (bottom), where yellow indicates higher dots density than blue. (**B**) Correlation between empirical and predicted regional HC degrees (top-left) and effect sizes of degrees (bottom-left); empirical and predicted regional maps of HC degrees (top-middle) and effect sizes of degrees (bottom-middle); and significant (*P*_spin_ < 0.05) PCA-CCA neurobiological loadings of HC degrees (top-right) and effect sizes of degrees, where data (*n* = 1,000 bootstrapped samples) are presented as mean values +/- standard deviation (bottom-right). (**C**) Regional associations between cognition and both MIND degree and centiles in SSD, where red indicates a positive correlation (left, top and bottom, respectively). Correlation between empirical and predicted regional MIND-cognition (top-middle) and centile-cognition (bottom-middle) associations in SSD, along with their empirical and predicted regional maps, where blue and red indicate a negative and positive correlation, respectively. Significant (*P*_spin_ < 0.05) PCA-CCA neurobiological loadings of MIND-cognition (top-right) and centile-cognition (bottom-right) associations in SSD. 5-HT_1A_, 5-HT_1B_, 5-HT_2A_, 5-HT_4_, 5-HT_6_, serotonin receptors; 5-HTT, serotonin transporter; Astro, astrocytes; CB_1_, cannabinoid receptor; CBF, cerebral blood flow; CBV, cerebral blood volume; CMRGlu, cerebral metabolic rate of glucose; CMRO_2_, cerebral metabolic rate of oxygen; D_1_, D_2_, dopamine receptors; DAT, dopamine transporter; Developmental exp., developmental expansion; Evolutionary exp., evolutionary expansion; Gene PC1, gene expression PC1; H_3_, histamine receptor; HC, healthy control; M_1_, acetylcholine receptor; Micro, microglia; MIND, Morphometric INverse Divergence; MOR, opioid receptor; Neuro-Ex, excitatory neurons; Neuro-In, inhibitory neurons; NMDA, glutamate receptor; OPC, oligodendrocyte precursor cells; Scaling NIH, allometric scaling from National Institutes of Health; Scaling PNC, allometric scaling from Philadelphia Neurodevelopmental Cohort; SSD, schizophrenia spectrum disorder; Thickness, cortical thickness; VAChT, acetylcholine transporter.

Finally, we performed two additional PCA-CCA analyses on the regional maps that represented the positive association between cognition and both MIND degree and centiles in SSD (Fig. 4C left, top and bottom, respectively). These maps indicated that lower cognition scores were associated with lower regional degrees and centiles in SSD. No significant associations were found between symptomatology and degree or centiles (see Supplementary Figs. 4 and 11). The PCA-CCA models were significant for both association maps of MIND (Fig. 4C top-middle; r = 0.52; *P*_spin_ = 0.001) and centiles (Fig. 4C bottom-middle; r = 0.51; *P*_spin_ = 0.002), where the predicted maps resembled their corresponding empirical values. Both models exhibited significant loadings (Fig. 4C right). Cortical expansion (Evolutionary and Developmental exp., Scaling PNC) and neurotransmitters (MOR, CB_1_, mGluR_5_, 5-HT_4_, 5-HT_2A_, 5-HT_1B_) demonstrated a significant positive contribution to the MIND-cognition association, indicating that the presence of these neurobiological features is highly co-localized with regions where lower cognition scores are associated with lower MIND degree. The features that positively accounted for explaining the regional associations between cognition and SSD centiles were cortical expansion (Evolutionary exp., Scaling NIH, Scaling PNC), neurotransmitters (5-HT_4_, 5-HT_2A_, CB_1_, M_1_, D_2_), and layer thickness (Layer I-III, V, and VI), indicating a high presence of these features in regions with lower centiles and cognition scores. As a sensitivity test, we performed Partial Least Square (PLS) analyses to assess whether the loading estimations were influenced by the PCA-CCA method used here. A high correlation (rLJ>LJ0.95) between loadings derived from both methods was found for significant models (Supplementary Fig. 21). Additionally, loadings remained consistent when we re-ran the PCA-CCA analyses with an explained variance of 60% to prevent overfitting (Supplementary Fig. 22).

## Discussion

In the present work, we investigated brain structure and organization by characterizing Morphometric Inverse Divergence (MIND) networks in individuals with schizophrenia spectrum disorders (SSD). We first assessed the differences between MIND networks of individuals with different SSD clinical status and healthy controls (HC). We additionally determined the regional connectivity to disease epicenters for each individual to assess how clinical status was influenced by the regions preferentially linked to atypical maturing areas. Finally, we explored how MIND organization in SSD relates to maturational events and neurobiological features.

Extensive evidence has shown that schizophrenia impacts both brain structure and connectivity^48^. In this context, recent morphometric studies have investigated individual structural similarity patterns to assess how the disease is associated with the expression of schizophrenia-related genes, leading to a widespread dysconnectivity pattern^6,49,50^. Here, we exploited the MIND method, identifying reductions in the cingulate, temporal, and insular lobes in individuals with SSD, indicating greater dissimilarity between these regions compared to HC. These findings align with previous research, providing new evidence on the dysconnectivity hypothesis of psychosis^6,49,51,52^. This dysconnectivity may result from the disintegration of nodes or hubs linked to structural changes in association fibers and aberrant control of synaptic plasticity, ultimately leading to abnormal functional connectivity in schizophrenia^51^. However, MIND increases were also found in regions connecting the frontal, parietal, and occipital lobes, indicating greater similarity between these regions, potentially reflecting increases in myelination that facilitates the trans-synaptic propagation of pathology^53^. To our knowledge, only one study has examined MIND alterations in individuals with early-onset schizophrenia, reporting significant changes in degree, though these were limited to a reduced number of regions^50^. However, studies in chronic schizophrenia have mainly reported increasing structural covariance strength as the disease progresses^54^, which might be attributed to disease progression, treatment effects, or medication use.

Abnormal connectivity has been proposed as a fundamental factor underlying cognitive deficits and clinical symptoms in patients with psychotic disorders^55^. In this study, individuals with more impaired general psychopathology showed reductions in edges connecting the temporal, insular, and cingulate lobes, while those with less impaired general psychopathology showed no significant differences. Accordingly, individuals with deeper cognitive impairment showed reductions in the temporal and cingulate lobes, while those with more preserved cognition did not. This could reflect a cognitive deficit associated to structural disconnections in psychosis-related individuals^56^. On the other hand, individuals with higher symptomatology reflected common nodal degree reductions in the rostral anterior cingulate region, which aligns with the notion that symptoms are associated with a reduction in thalamocortical connectivity to the middle frontal gyrus, anterior cingulate, and insula regions^55^.

Extensive research supports the notion that psychiatric disorders originate from alterations in specific brain regions, known as epicenters, which then propagate to other areas through existing connectivity patterns^27,57^, involving genetic, trans-neuronal, structural and functional processes^16,28^. This model aligns with findings in neurological conditions, where epicenter frameworks based on structural and functional connectivity and network diffusion models have successfully predicted the progression of neurodegenerative changes^58–60^. One proposed mechanism for network-specific degeneration is that axonal connectivity (connectome) guides the spread of misfolded proteins into new regions^58,61^, leading to cell death and atrophy^28^. Alternatively, dysfunction in one region may trigger abnormal activity in connected areas, which, over time, could lead to structural changes as a result of disrupted neurotransmission or diminished trophic support^7,59,60^. A further possibility is that genetic variants of genes involved in schizophrenia, such as G72 and DISC1, may modulate regional changes during SSD progression^62,63^. Here, although MIND degree was not associated with SSD epicenters (centiles), we reported that MIND edges were stronger (i.e., higher structural regional resemblance) between pair of regions exhibiting similar co-vulnerability to psychosis^37^. This widespread distribution of regions showing significant connectivity to disease epicenters likely reflects the globally distributed nature of schizophrenia, rather than being indicative of focal regional abnormalities^64^. Given this broad network-level disruption, even moderate alignment across widely distributed regions can yield a large number of statistically significant associations. This finding cannot be attributed to a specific permutation method, as both Moran spectral randomization, which accounts for spatial autocorrelation^65^, and a rewired null model that preserves the empirical degree sequence and edge length distribution^66^ produced similar results.

Maturation of higher-order cognitive and affective functions is disrupted in schizophrenia^67^. Here, while centile decreases affected lower-order primary regions associated with sensorimotor functions (as observed in antipsychotic-naïve first-episode psychosis individuals)^68^, connectivity reductions impacted higher-order association areas linked to associative functions^46^. Accordingly, the hierarchical organization of the cortex has also been demonstrated to be altered in different fMRI studies of schizophrenia, affecting the secondary visual-to-sensorimotor axis rather than the primary sensorimotor-to-association axis^40,69^. Taken together, these findings suggest that SSD-related impairments target specific hierarchical processes, where focal disruptions in somatosensory-motor areas may extend into higher-order regions^69^.

Understanding the interaction between brain maturation and abnormal structural connectivity is essential for gaining deeper insights into the neurodevelopmental origins of psychiatric conditions^70,71^. Here, we found that regions maturing earlier tend to exhibit higher MIND degree, aligning with studies indicating that rapidly maturing hubs are located in high-level functional networks, potentially facilitating their functional and anatomical maturation^72^. Additionally, the significant negative association between SSD degree effect sizes and both volume and velocity age peaks indicated that late-maturing regions are particularly affected by the disease. This aligns with fMRI connectivity and DWI studies of schizophrenia^73,74^, which suggest that dysmaturation of neural connectivity is a key component of its pathophysiology^75^. We also found that local atypical maturation, as indicated by centile analyses, is more prominent in early-maturing regions, consistent with the previously observed lower hierarchical role of these regions. Altogether, these findings suggest a complex interaction between local and network-level SSD alterations, cortical processing hierarchy, and maturational events.

Recent research has co-localized maturational events and structural disease-related alterations with the underlying neurobiology^26^. Particularly, abnormal patterns of MIND connectivity in major depressive disorder have been associated to the cannabinoid receptor and the brain-wide expression of a genetic combination^76^. In this study, we identified a common pattern of neurobiological features across connected regions, suggesting that the macroscopic structural properties used to estimate MIND edges are heavily influenced by underlying microstructural organization^77^, neurotransmitter systems^43^, and metabolism^78^. This finding aligns with the higher gene co-expression found in highly MIND-connected regions^15^. However, the differences between SSD and HC in MIND edges were not linked to overall shared neurobiology, but rather with specific neurobiological types that were either co-localized or anti-co-localized with differences in SSD edges. At the regional level, the positive contribution of excitatory and inhibitory neurons to MIND edges based on CCA loading estimates provided further evidence of the fundamental role that neuron density plays in shaping the architecture and connectivity of the primate cerebral cortex^79^. The high presence of OPC, microglia, and astrocytes in weakly-connected regions aligns with the role these cell types play in actively guiding brain development^80–83^. Furthermore, these glial cells also contributed to explaining the reduction in MIND degree of patients with SSD. This disruption in astrocyte differentiation may lead to abnormal glial coverage and hypomyelination, resulting in insufficient synaptic support, compromised white matter integrity, and transmission desynchronization, ultimately causing network dysconnectivity^84^. Neurotransmitters and layer thickness also contributed to these alterations in MIND degree due to SSD. The hypothesis of anatomical disconnection in schizophrenia has also been linked to synaptic damage, as well as dysfunction of dopaminergic, cholinergic, or glutamatergic systems^85^. A principal component of cortical gene expression architecture has been found to be specifically associated with marker genes of layers II and III, and supragranular thinning in schizophrenia^86^.

A low presence of myelin and cerebral blood volume was related to regions where lower MIND degrees were associated with lower cognition scores, aligning with the disrupted relationship found between myelination and neurocognition in patients with schizophrenia^87^. Chronic low cerebral blood volume in schizophrenia may lead to neural hypoactivation, ultimately disrupting cortical cytoarchitectural organization and potentially contributing to brain atrophy and associated cognitive impairments^88^. However, a high presence of neurotransmitters and cortical expansion features were associated with regions where both lower MIND degrees and centiles were associated with lower cognition scores, which aligns with the previous studies relating the imbalance in glutamate, acetylcholine, GABA, dopamine, and histamine with cognitive deficits in patients with schizophrenia^89^. Thus, MIND allows for a more precise and personalized biologically grounded representation of an individual’s brain architectome, potentially serving as a biomarker for diagnostic and stratification purposes, the identification of biologically defined subtypes, and the development of more targeted and effective therapeutic strategies.

The present findings must be interpreted with several considerations. First, MRI data were obtained from different 1.5 and 3T MRI scanners. Nevertheless, site harmonization was performed to MIND networks, after which MIND degree was highly correlated between scanners (r = 0.96; Supplementary Fig. 1). Second, the current implementation of MIND networks is limited to vertex-level data derived from MRI-based cortical surface reconstructions^15^. As a result, subcortical structures were excluded from this study, despite their well-established clinical relevance. Third, MIND networks derived from cross-sectional data were associated with dynamic processes of maturation. Analyzing longitudinal trajectories of MIND changes would offer a more precise understanding of how maturational processes interact with disease progression and treatment effects. Fourth, given the limited sample size scenario (68 regions), the CCA multivariate approach has a risk of over-fitting^90^. To address this issue, we first applied PCA prior to CCA. Fifth, the maturational events and neurobiological data used here were derived from normative non-psychotic individuals, which limits the capacity to capture patient-specific variability. A recent PET study, however, has demonstrated that reduced synaptic density in schizophrenia is also spatially aligned with altered normative distributions of neurotransmitter levels, suggesting that the synaptic deficits are shaped by the molecular similarity between regions^91^. Nevertheless, relying on external cellular and molecular datasets limits our ability to capture the biological attributes underlying between-participant variability in brain structure. In this context, a recent study that combined antemortem neuroimaging, molecular, and genetic data with postmortem dendritic spine density and morphometric attributes demonstrated that modules of synaptic protein and expression can explain individual differences in both functional connectivity and structural covariation^92^. These findings highlight the feasibility of cross-scale data integration to enhance our understanding of brain connectivity. Future work with neurobiological data of individuals with SSD would provide valuable insights into the molecular alterations causally associated with abnormal brain alterations.

In summary, using MIND networks, we identified reductions in regional structural similarity within the temporal, cingulate, and insular lobes of individuals with SSD, alongside an increase in the occipital lobe. These decreases in MIND connectivity were particularly pronounced in individuals with more severe clinical status. Moreover, these MIND reductions were co-localized with maturational events and were more prominent in regions characterized by a high normative presence of neurotransmitters, specific cortical layer thickness, and astrocytes, as well as decreased microstructure and metabolism. Altogether, these findings contribute to a better understanding of the disease process, highlighting the role of brain structure, maturational events, and neurobiology in shaping clinical status during the early stages of the disease.

## Methods

### Subjects

Magnetic Resonance Imaging (MRI) data from individuals who were determined to have experienced a first-episode non-affective psychosis, confirming the presence of a schizophrenia spectrum disorder (SSD; *n* = 352; *n*_female_ = 138; *age* = 31.43 ± 8.78), including schizophrenia (*n* = 169), schizophreniform (*n* = 102), brief psychotic (*n* = 49), unspecified psychotic (*n* = 26), and schizoaffective (*n* = 6) disorders, along with a group of healthy controls (HC; *n* = 195; *n*_female_ = 75; *age* = 30.64 ± 7.66), were obtained from *Programa de Atención a las Fases Iniciales de Psicosis* (PAFIP)^93^. Individuals with SSD had no prior antipsychotic medication or were minimally medicated. All diagnoses were made by an experienced psychiatrist using the Structured Clinical Interview for DSM-IV (SCID-I), confirming the presence of schizophrenia or other psychotic disorder after 6 months of the first visit. Participants who met criteria and provided written informed consent were entered into PAFIP. Additional inclusion and exclusion criteria can be found in *Supplementary Subjects and Methods*.

This program has been approved by the Ethics Committee for Clinical Research (CEIC - Cantabria) in accordance with international standards (NCT0235832 and NCT02534363 clinical trial numbers).

### MRI acquisition, parcellation, and volume extraction

149 SSD and 80 HC MRI scans were obtained using a 1.5 T General Electric SIGNA System (GE Medical Systems, Milwaukee, WI), and 203 SSD and 115 HC scans using a 3 T Philips Medical Systems MRI scanner (Achieva, Best, The Netherlands) at the Hospital Universitario Marqués de Valdecilla (HUMV). The parameters for 1.5 T were: TE = 5 ms, TR =24 ms, NEX = 2, rotation angle = 45°, FOV = 26×19.5 cm, slice thickness = 1.5 mm, and a matrix of 256×192; whereas for 3 T were: TE = 3.7 ms, TR = 8.2 ms, flip angle = 8°, acquisition matrix = 256×256, voxel size = 0.94×0.94×1 mm and 160 contiguous slices. T_1_-weighted images were first visually inspected for artefacts and gross anatomical abnormalities, and were processed using FreeSurfer 6.0.0 (http://surfer.nmr.mgh.harvard.edu) applying the recon-all pipeline to enhance gray-white matter boundary delineation.

Cortical parcellation was performed using Desikan-Killiany atlas, and volumetric measurements were derived for each of the 68 region-of-interest. Quality control procedures were implemented to ensure the accuracy and reliability of the cerebral volumes derived. Thirteen HC subjects exhibiting visual artifacts or processing errors were excluded from further analysis. No further participants were excluded, as their global degree fell within the range of the average global degree ± 5 standard deviations (0.166 ± 0.034).

### MIND edges and degrees estimation

For each subject, a Morphometric INverse Divergence (MIND) network was constructed by computing the Kullback-Leibler (or Jeffrey’s) divergence of multiple MRI features (cortical thickness, mean curvature, sulcal depth, surface area, and gray matter volume) between each pair of regions^15^. Thus, each cortical region was characterized by a multidimensional distribution of these features measured at sub-voxel vertex resolution on a cortical surface mesh. To calculate the multivariate divergence between pairs of regions directly from the observed vertex-level data, this method leverages a *k*-nearest neighbor approach, where a decrease in the divergence of these multivariate distributions (or an increase in their similarity) indicated an increase in MIND. This resulted in a 68×68 edge matrix (bounded between 0 and 1), representing the structural similarity between regions (also referred here as structural connectivity). To correct site-related batch effects while preserving age, sex, and psychosis-related effects including sex-psychosis interaction, we used ComBatLS^94^, a recent extension of ComBat method that preserves biological variance, while reducing sex biases. To regress the effect of age, sex, and the estimated total intracranial volume (eTIV), we analyzed the residuals (rather than the raw MIND networks) derived from the linear model shown in Equation (1).

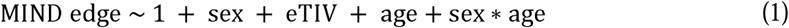

All non-binary variables were z-scored prior to regression. Individual MIND nodal degrees were computed by averaging their corresponding MIND residuals (referred here as edges), resulting in an average within-subject similarity degree for each region. Thus, while MIND edges represent how structurally similar each pair of regions is, MIND degrees reflect the average structural similarity of a given region with the rest of the brain.

MIND edges and degrees of patients with SSD were compared to HC by calculating the effect size (Cohen’s d) between each SSD and HC edge and region, respectively. To statistically assess these differences, a permutation test was applied by randomly shuffling group assignment to create a null distribution (10,000 permutations). Additionally, tail area-based false discovery rate (FDR) correction^95^ across brain regions and edges was applied to account for multiple comparisons (with an alpha set at < 0.05).

### Assessment of cognition and symptomatology

Effect sizes of MIND edges and degrees were stratified based on the general psychopathology, the average global cognitive function (i.e., cognition), and the clinical symptomatology of drug-naïve first-episode psychosis (FEP) individuals. Overall cognitive score was assessed for the following domains: (1) verbal memory: Rey Auditory Verbal Learning Test (RAVLT) long-term recall score; (2) visual memory: Rey Complex Figure test (RCFT) long-term recall score; (3) motor dexterity: grooved pegboard (GP), time to complete with dominant hand; (4) executive functions: Trail Making Test part B (TMTb); (5) working memory: WAIS III-Backward Digits (BD) total score; (6) speed of processing: WAIS III-Digit Symbol (DS) standard total score; (7) attention: Continuous Performance Test Degraded-Stimulus (CPT-DS), the total number of correct responses^96^. Symptomatology was assessed with the Brief Psychiatric Rating Scale (BPRS) (expanded version of 24 items), and the clinical symptoms of psychosis were assessed with the Scale for the Assessment of Positive Symptoms (SAPS) and the Scale for the Assessment of Negative Symptoms (SANS). General psychopathology was assessed by performing a *k*-means clustering based on the different scales considered (global cognitive function, BPRS, SAPS, and SANS), stratifying individuals as having a ‘good’ or ‘poor’ general psychopathological status. Individuals with SSD with a ‘poor’ status in cognition or symptomatology were defined as those scoring below the median score for cognition and at or above the median for symptomatology (cognition, *n* = 151; BPRS, *n* = 165; SAPS, *n* = 168; SANS, *n* = 168; see Supplementary Fig. 3 for mean and median values across scales). Conversely, individuals with a ‘good’ status were those who had scores at or above the median score for cognition and below the median for symptomatology.

### Computing connectivity to disease epicenters

Regional centiles were computed for each participant by benchmarking regional MRI volumes against previously established normative trajectories (from approximately 120,000 participants), which adjusts for age-, sex-, and site-effects^36^. ComBatLS was also applied to centiles to correct for site-effects. SSD centile distributions were first compared to HC using the Wilcoxon rank-sum test (FDR-corrected) and subsequently correlated with the average regional structural similarity (nodal degree) of HC individuals to assess whether a stronger structural connectivity was associated with atypically maturating regions in SSD. The structural co-vulnerability to psychosis matrix (34×34 averaged across hemispheres), derived in our recent study^37^, was constructed by correlating the regional effect sizes of the centiles across different psychosis-related groups, including first-degree relatives of patients with schizophrenia or schizoaffective disorder, individuals who had psychotic experiences, individuals from this cohort (SSD), and patients with chronic schizophrenia or schizoaffective disorder (see Supplementary Table 1 for demographic details of these cohorts).

Regions with the most pronounced morphological alterations at disease onset (i.e., most reduced centiles in SSD) were defined as disease epicenters^16,28^. For each individual with SSD, the connectivity to disease epicenters for each region was computed as the Pearson correlation *r* between the SSD edge of that region and the average regional centiles in SSD. Consequently, regions that were preferentially associated with reduced centiles were considered connected to disease epicenters. For each region, the *P*-value (tail area-based FDR-corrected) was computed as the proportion of permuted *r*-values, averaged across individuals, that were more extreme than the observed average values. The null distributions of *r*-values were derived from a rewiring null model (Betzel) that preserves the exact degree sequence and edge weight distribution, while also approximates its edge length distribution (based on geodesic distance between regions) and the relationship between edge weights and lengths^66^. Moran spectral randomization, a method for generating spatially constrained null models that accounts for spatial dependencies^65^, was additionally employed.

As a sensitivity analysis, connections to epicenters were computed for HC subjects correlating the HC edges of each region with the average regional HC centiles under the assumption that no preferential connectivity with ‘healthy’ epicenters would be revealed.

### MIND association with cortical maturation

Centiles and effect sizes were associated with a sensorimotor-association regional ranking reported by Sydnor *et al*.^46^, which ranges from lower-order sensorimotor to higher-order associative functions. Two additional maturational features derived from the same normative trajectories as centiles were considered: volume peak, representing the age at which regional volume reaches its highest point; and velocity peak, indicating the age at which regional volume growth is the fastest. These features were compared to MIND networks (both HC and effect sizes) and SSD centiles. All regional-based associations, computed after accounting for regional volumes, were statistically assessed by applying a spin test (*n* = 10,000) where spatial autocorrelation was preserved (see *Supplementary Subjects and Methods* for details). The Betzel rewiring null model preserving the degree sequence and approximately the edge length distribution, as well as Moran spectral randomization, were employed on the MIND edge-based associations to account for basic network properties and spatial dependencies, respectively.

### Identifying neurobiological features associated with MIND networks stratified by clinical status

The spatial maps of 46 molecular and micro-architectural features, including neurotransmitters, cell types, layer thickness, microstructure, cortical expansion, and metabolism, were collected across multiple studies (see Supplementary Table 2 for a complete list of neurobiological features, and refs. ^42,97,98^ for population-specific characteristics)^47^. These maps were obtained in surface or volumetric spaces using *neuromaps* toolbox, available at https://github.com/netneurolab/neuromaps, and were parcellated according to Desikan-Killiany atlas. In cases where multiple maps were available for a single feature, a weighted average was taken. The neurobiological similarity matrix was computed by correlating the 46 spatial maps between regions. We assessed whether the neurobiological similarity was co-localized with MIND edges using both the degree- and edge length-preserving null model, as well as Moran spectral randomization.

Canonical Correlation Analysis (CCA) is a powerful multivariate method for capturing associations between two modalities of data (e.g., brain and behavior; see *Supplementary Subjects and Methods* for details)^99^. A combined Principal Component Analysis (PCA) and CCA approach was employed to investigate the regional associations of MIND degree with the neurobiological features. The contribution of each feature to this neurobiology-MIND association was calculated by identifying the model loadings^99^. Statistical significance of the models and loadings was assessed using the spin test.

To identify the neurobiological features associated with phenotypes that were linked to clinical status, we performed PCA-CCA analyses on the regional maps that represented the associations between clinical status and both MIND degrees and centiles. Since symptomatology did not show any significant association with MIND or centiles (see Supplementary Figs. 4 and 11), we did not assess the neurobiological association with these maps.

## Data availability

According to the informed consent, MRI data from PAFIP dataset is available from the corresponding author on request. All maturational features, including centiles and the volume and velocity peaks, were obtained from https://github.com/brainchart/Lifespan. The neurobiological maps were obtained using *neuromaps* toolbox,^97^ available at https://github.com/netneurolab/neuromaps. The Desikan–Killiany parcellation atlas used for the neurobiological mapping was obtained from netneurotools (https://github.com/netneurolab/netneurotools). Source data are provided with this paper.

## Code availability

All code used to perform the analyses can be found at https://github.com/NeuroimagingBrainNetworks/NeurobiologyMINDPsychosis and https://doi.org/10.5281/zenodo.16090993. The code used for computing MIND networks can be found at https://github.com/isebenius/MIND. The code used for the PCA-CCA analyses is available at https://github.com/NeuroimagingBrainNetworks/cca_pls_toolkit.

## Acknowledgments

R.R.G. is funded by *Plan de Generación de Conocimiento* (PID2021-122853OA-I00), *Plan de Consolidación* (CNS2023-143647) and *ERANET Neuron JTC* 2023 (ERP-2023-23684211). Both R.R.G. and N.G.S. are funded by the *EMERGIA Junta de Andalucía* program (EMERGIA20_00139). R.A.A. is funded by *Plan de Consolidación* (CNS2022-136110).

## Author Contributions

N.G.S. performed data curation, methodological design, data analysis, and drafted the manuscript; R.A.I.B., P.S., A.M., J.Se., I.S., C.A.M., L.D., G.S., S.E.M., M.R.V., R.A.A., J.V.B., B.M., J.Su., B.C.F., and R.R.G contributed to data acquisition, provided advice on data analysis, and participated in writing and editing the manuscript. R.R.G. also contributed to conceptualization and supervision of the work. All authors approved the submitted version of the manuscript.

