## Supplemental Material for "Reduced brain structural similarity is associated with maturation, neurobiological features, and clinical status in schizophrenia"

**This PDF file includes:**

**Other Supplementary Information for this manuscript includes:**

Source Data (separate .xlsx file)

### Supplementary Methods

#### Dataset description

Subjects from *Programa de Atención a las Fases Iniciales de Psicosis* (PAFIP) dataset were screened for the following criteria[1,2]: (1) age 15 to 60 years; (2) DSM-IV criteria for a principal diagnosis of schizophreniform disorder, schizophrenia, schizoaffective disorder, brief reactive psychosis, schizotypal personality disorder, or psychosis not otherwise specified; (3) habitually living in the catchment area; (4) no prior treatment with antipsychotic medication or, if previously treated, a total lifetime of adequate antipsychotic treatment of less than 6 weeks. Patients were excluded for any of the following reasons: 1) met the DSM-IV criteria for drug dependence (except nicotine dependence), 2) met the DSM-IV criteria for mental retardation, or 3) had a history of neurological disease or head injury. There were no additional exclusion criteria for MRI except those specific to scanning logistics (e.g., claustrophobia, braces). The healthy volunteer group was recruited from the community through advertisements. They were required to have no current or previous psychiatric, mental retardation, neurological or general medical illnesses, including substance dependence and significant loss of consciousness, as determined by using an abbreviated version of the Comprehensive Assessment of Symptoms and History (CASH)[3]. They were selected to have a similar distribution in age, gender, and drug use history to the patients. Clinical records and family interviews also confirmed the absence of psychosis in first-degree relatives. After a detailed description of the study, each subject gave written informed consent to participate.

#### Spin test

The spin test projects brain regions to a sphere using spherical coordinates generated during cortical-surface extraction. These spherical projections of brain annotation maps are rotated to randomize the relationship between cortical attributes and annotations[4,5]. Each coordinate is assigned to its nearest rotated counterpart, resulting in a map where spatial autocorrelation is preserved, while the correspondence between parcels and annotations is randomized. Parcels that were closest to the medial wall were assigned the value of the nearest neighboring parcel instead. This procedure was performed at parcel resolution, rather than the vertex resolution, to prevent data upsampling, and it was repeated 10,000 times to generate a parcellation-specific rotation matrix (available at https://github.com/frantisekvasa/rotate_parcellation).

#### Principal Component Analysis - Canonical Correlation Analysis (PCA-CCA)

A combined Principal Component Analysis (PCA) and Canonical Correlation Analysis (CCA) approach was employed to relate neurobiological maps to regional Morphometric Inverse Divergence (MIND) degrees and effect sizes. However, under scenarios where the sample size (68 cortical regions) is similar to or smaller than the number of variables (46 neurobiological features), standard multivariate models may overfit. To address this issue, a prior dimensionality reduction with PCA was performed with we *PCAtools* to determine the optimum number of PCs to retain. Elbow method and Horn's parallel analysis[6,7] provided a number of PCs of 6 and 5, respectively, which corresponded to an explained variance of 80% (Supplementary Fig. 17). To assess the statistical significance of the models, the spatial autocorrelation-preserving permutation spin test was used.

The PCA-CCA models identified a set of weights ($w_x$) that the resulting linear combination (weighted sum) of the neurobiological maps constituted a set of predicted degrees and effect sizes that are, by construction, correlated with the empirical values. To assess the significance of the model weights, the autocorrelation-preserving spin test was employed. To estimate the standard errors, 1,000 bootstrap samples were created by sampling with replacement the observations of the molecular maps. Finally, to ascertain the extent to which each neurobiological feature contributed to the predicted values, a set of loadings was computed as the Pearson correlation between the neurobiological maps and the dimensionally-reduced predicted degrees or effect sizes.

### Supplemental Figures

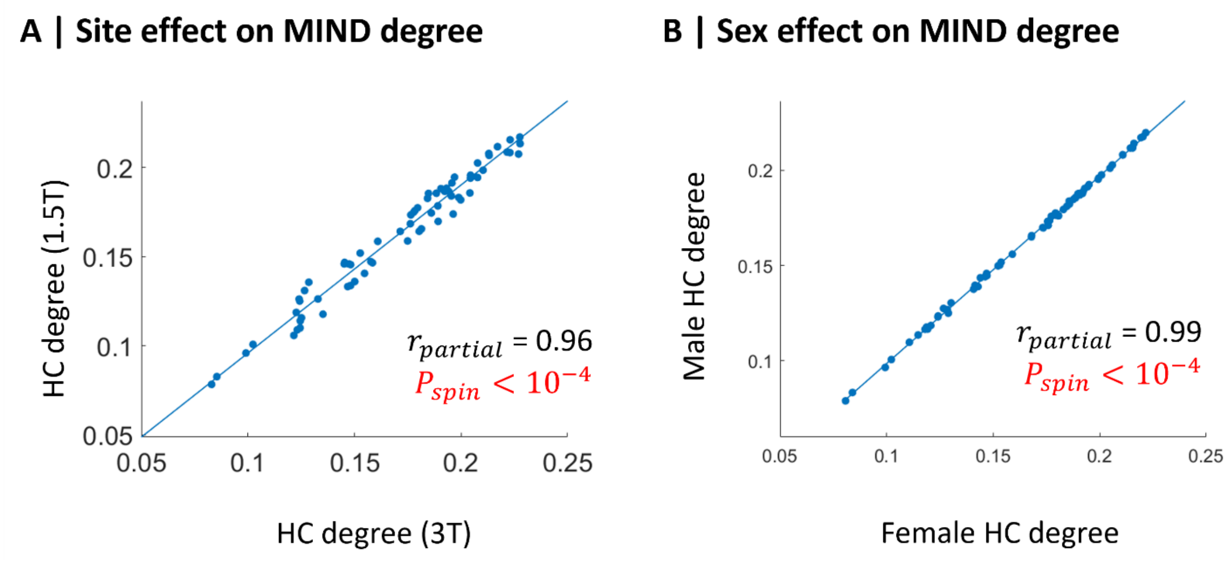

**Supplementary Fig. 1** Association between HC MIND degrees assessed (**A**) with different scanners (1.5 and 3T); and (**B**) for males and females (two-sided test). HC, healthy control; MIND, Morphometric INverse Divergence.

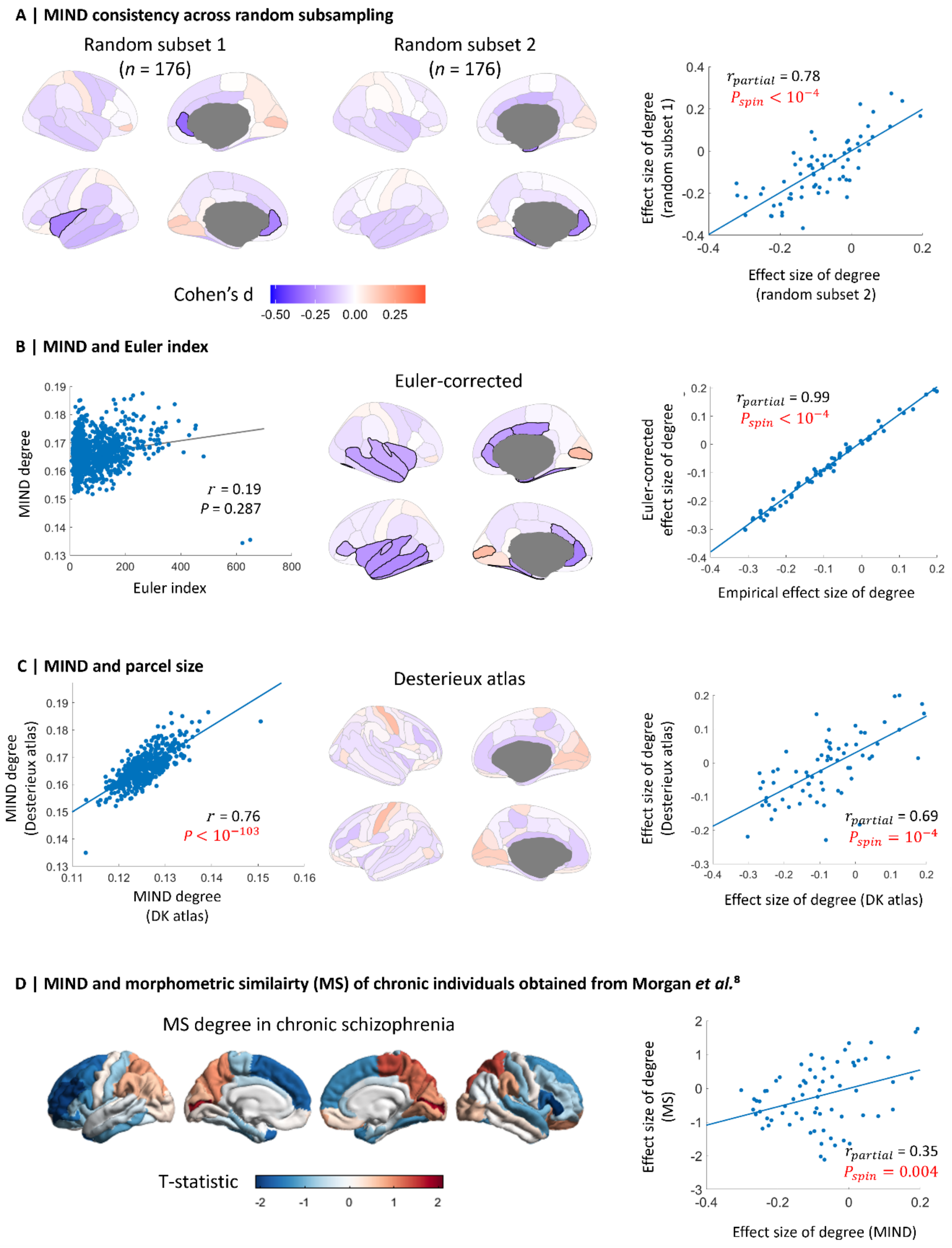

**Supplementary Fig. 2** (**A**) Regional effect sizes (Cohen's d) computed between each randomly selected half of the sample of SSD patients ($n$ = 176) and HC MIND degrees (left and middle). Blue and red indicate a significant reduction and increase of MIND degree in SSD, respectively. Association between regional effect sizes of MIND degree of each subset (right; two-sided test). (**B**) Association between global MIND degree and the Euler index (left). Regional effect sizes computed between Euler-corrected SSD and HC MIND degrees (middle). Association between Euler-corrected regional effect sizes of MIND degree and the empirical effect sizes of MIND degree (right; two-sided test). (**C**) Association between global MIND degrees using DK and Desterieux atlases (left). Regional effect sizes computed between SSD and HC MIND degrees using Desterieux atlas (middle). Association between regional effect sizes of MIND degree using DK and Desterieux atlases (right; two-sided test). (**D**) Regional effect sizes of MS degree derived from chronic schizophrenia individuals in Morgan *et al.*[8] (left). Association between regional effect sizes of MIND degree of SSD patients and MS degree of chronic patients (right; two-sided test). The highlighted nodes show those that exhibit significant differences from HC after tail area-based false discovery rate (FDR) correction (permutation test, $P < 0.05$). DK, Desikan-Killiany; MIND, Morphometric INverse Divergence; MS, morphometric similarity; SSD, schizophrenia spectrum disorder.

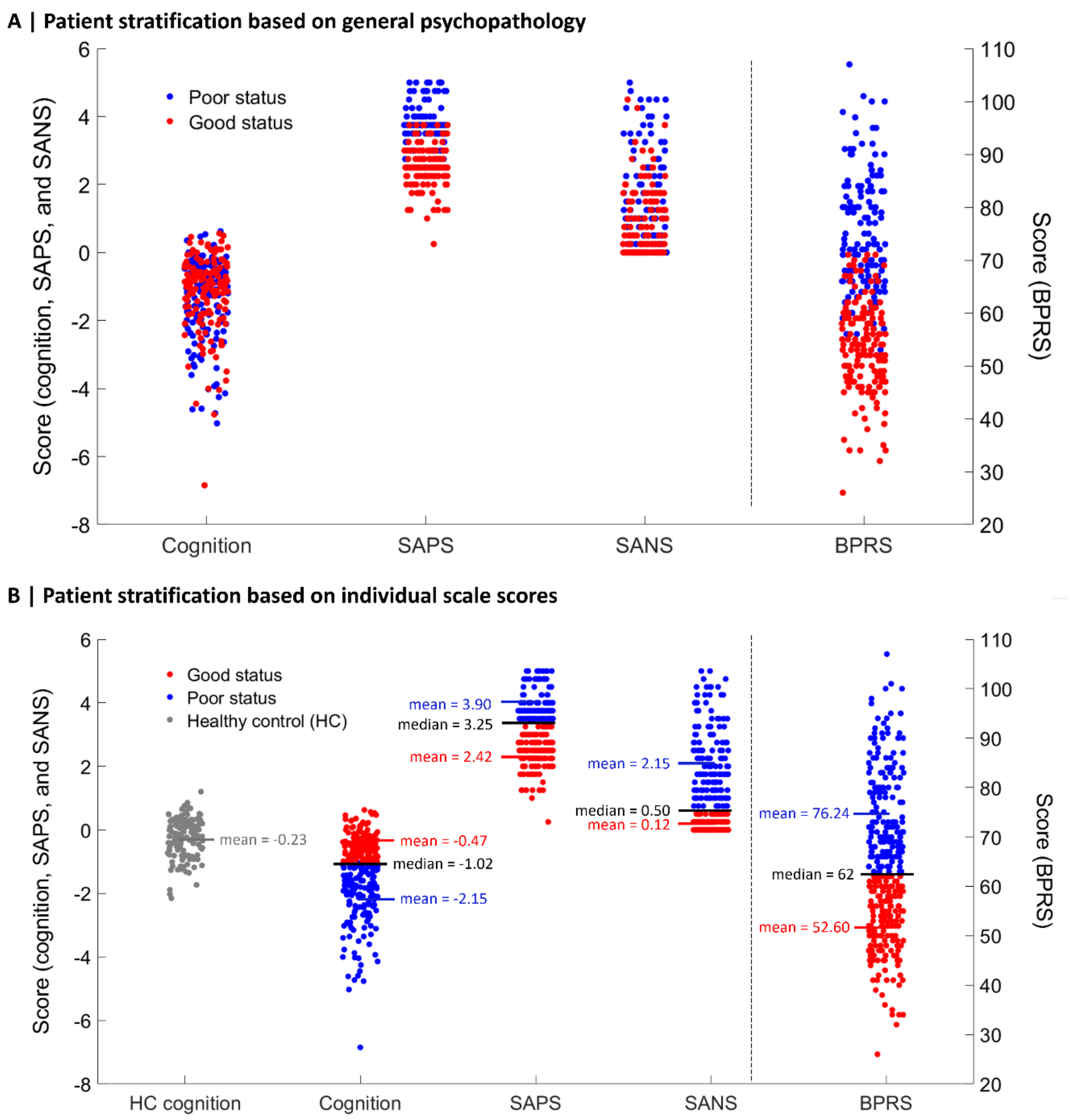

**Supplementary Fig. 3 Patient stratification according to their clinical status.** (**A**) Individual global scores of cognitive functioning and clinical symptomatology classified as 'poor' or 'good' status based on their general psychopathology, which was assessed through a *k*-means clustering performed across the different scales: global cognitive functioning (cognition), Brief Psychiatric Rating Scale (BPRS), Scale for the Assessment of Positive Symptoms (SAPS), Scale for the Assessment of Negative Symptoms (SANS). (**B**)

Individual global scores of cognitive functioning and clinical symptomatology stratified as 'poor' status if its value was below (cognition) or at or above (BPRS, SAPS, SANS) the median score of each scale; or as 'good' status if its value was at or above (cognition) or below (BPRS, SAPS, SANS) the median score.

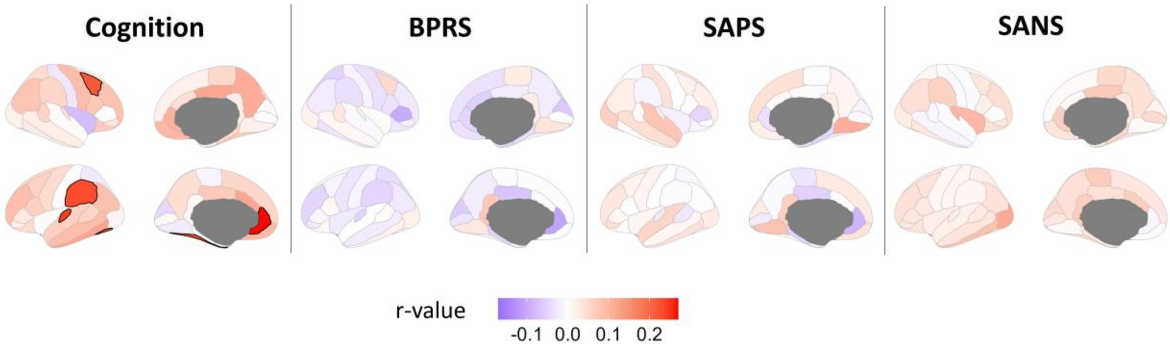

**Supplementary Fig. 4 Regional correlations between individual MIND degrees and their clinical status.** Blue and red indicate a negative and positive correlation, respectively. BPRS, Brief Psychiatric Rating Scale; SANS, Scale for the Assessment of Negative Symptoms; SAPS, Scale for the Assessment of Positive Symptoms.

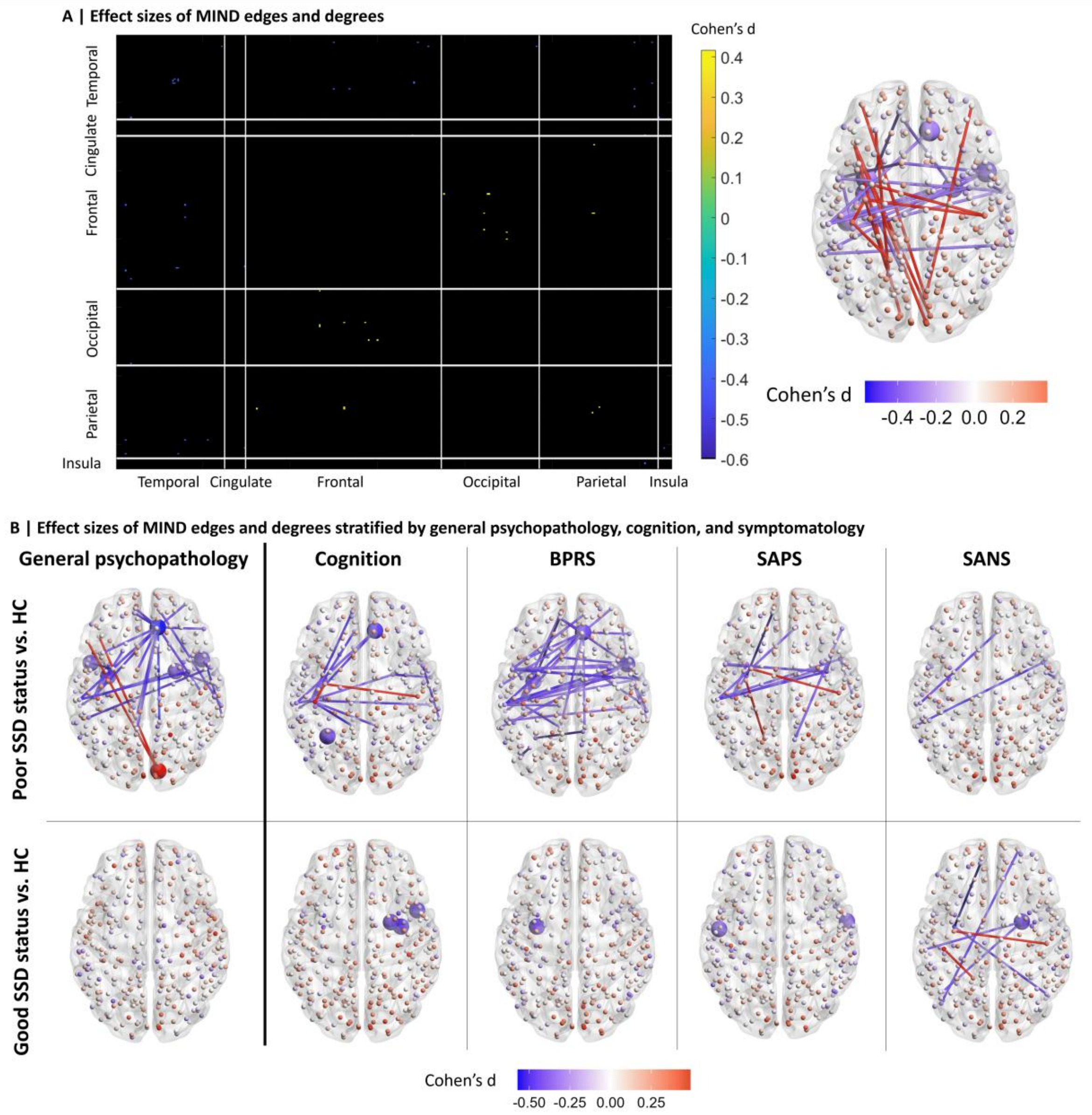

**Supplementary Fig. 5 MIND connectivity (DK-318-parcellated) in relation to clinical status.** (**A**) Effect sizes (Cohen's d) computed between SSD and HC MIND edges (left), where blue indicates a significant reduction of edge and degree in SSD, yellow and red indicate a significant increase, and black indicates no significant differences. Spatial 3D representation of significantly different edges and degrees in SSD compared to HC (right). (**B**) Effect sizes of MIND edges and degrees stratified by general psychopathology, cognition, and symptomatology. The Cohen's d of MIND degree for those individuals with SSD whose general psychopathology is more impaired, whose cognition is lower, and whose symptomatology (BPRS, SAPS, and SANS) is equal or higher than the median of the sample (top row). The Cohen's d for those whose general psychopathology is less impaired, whose symptomatology is lower, and whose cognition is equal or higher than the median of the sample is shown in the bottom row. The highlighted nodes and edges show those that exhibit significant differences from HC after tail area-based FDR correction (permutation test, $P < 0.05$). BPRS, Brief Psychiatric Rating Scale; HC, healthy control; MIND, Morphometric INverse Divergence; SANS, Scale for the Assessment of Negative Symptoms; SAPS, Scale for the Assessment of Positive Symptoms; SSD, schizophrenia spectrum disorder.

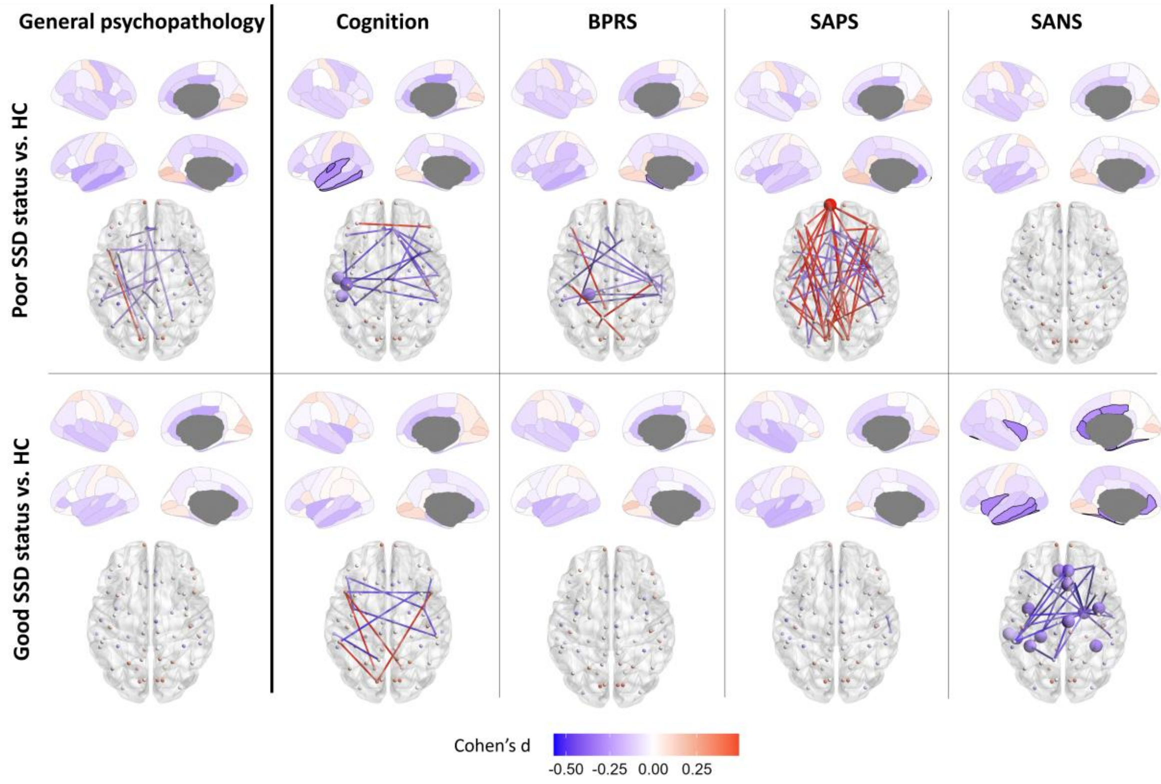

**Supplementary Fig. 6 Distinct effect sizes of MIND edges and degrees of each scale.** Distinct effect sizes of MIND edges and degrees stratified by general psychopathology, cognition, and symptomatology, where blue indicates a significant reduction of edge and degree in SSD, and red indicates a significant increase. The Cohen's d of MIND degree for those individuals with SSD whose general psychopathology is more impaired, whose cognition is lower, and whose symptomatology (BPRS, SAPS, and SANS) is equal or higher than the median of the sample (top row). The Cohen's d for those whose general psychopathology is less impaired, whose symptomatology is lower, and whose cognition is equal or higher than the median of the sample is shown in the bottom row. The highlighted regions show those regional degrees that exhibit significant differences from HC after tail area-based FDR correction (permutation test, $P < 0.05$). BPRS, Brief Psychiatric Rating Scale; HC, healthy control; SANS, Scale for the Assessment of Negative Symptoms; SAPS, Scale for the Assessment of Positive Symptoms; SSD, schizophrenia spectrum disorder.

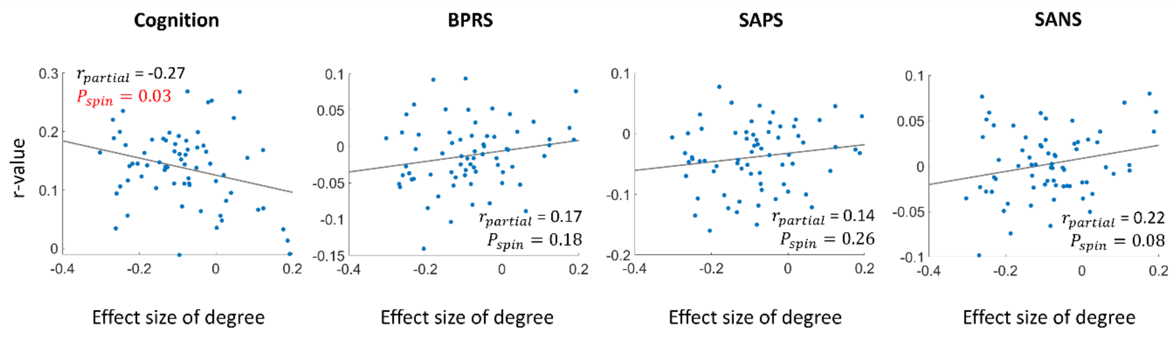

**Supplementary Fig. 7** Association (two-sided test) between the regional effect sizes of MIND degree and the regional r-values between MIND degree and the scores of each scale (cognition, BPRS, SAPS, and SANS). BPRS, Brief Psychiatric Rating Scale; SANS, Scale for the Assessment of Negative Symptoms; SAPS, Scale for the Assessment of Positive Symptoms.

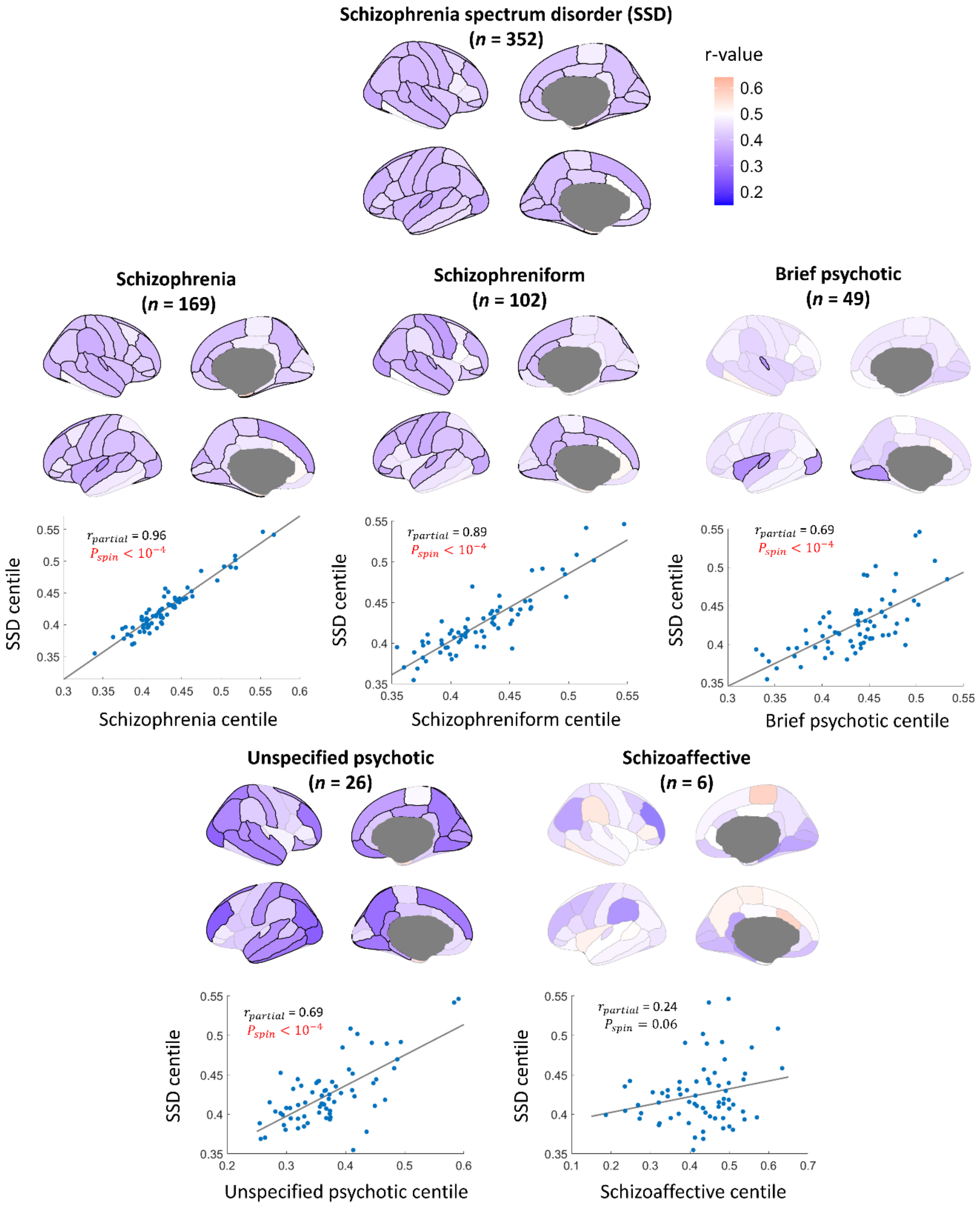

**Supplementary Fig. 8 Regional centiles for each SSD diagnostic group**. Regional centile maps for each SSD diagnostic group (brain plots), and its association with the average SSD centiles (scatter plots; two-sided test).

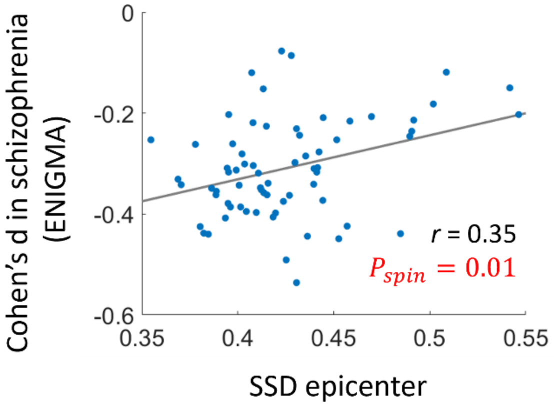

**Supplementary Fig. 9 Association (two-sided test) between SSD epicenters (i.e., centiles) and cortical thickness abnormalities in schizophrenia derived from ENIGMA[9].** Cortical thickness abnormalities were computed as the Cohen's d between chronic schizophrenia patients and controls. SSD, schizophrenia spectrum disorder.

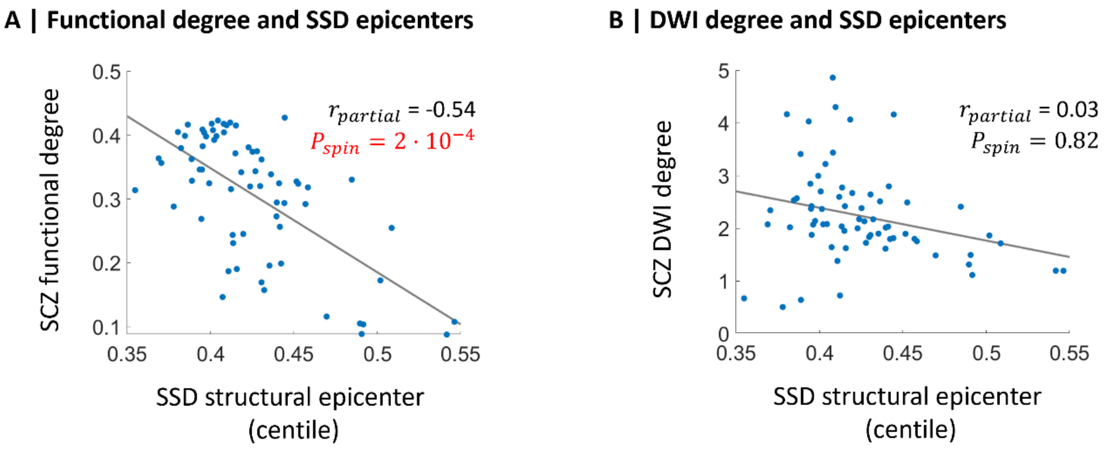

**Supplementary Fig. 10 Association (two-sided test) between SSD structural epicenters and the average connectivity in SCZ.** (**A**) Functional and (**B**) DWI degree derived from an ENIGMA schizophrenia study[10] were correlated with structural SSD epicenters. DWI, diffusion-weighted imaging; SCZ, schizophrenia; SSD, schizophrenia spectrum disorder.

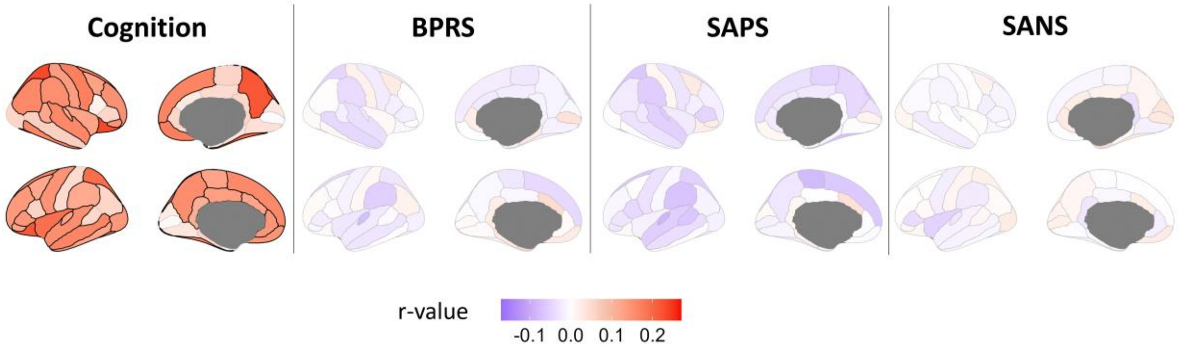

**Supplementary Fig. 11 Regional correlations between individual disease epicenters and their clinical status.** Blue and red indicate a negative and positive correlation, respectively. BPRS, Brief Psychiatric Rating Scale; SANS, Scale for the Assessment of Negative Symptoms; SAPS, Scale for the Assessment of Positive Symptoms.

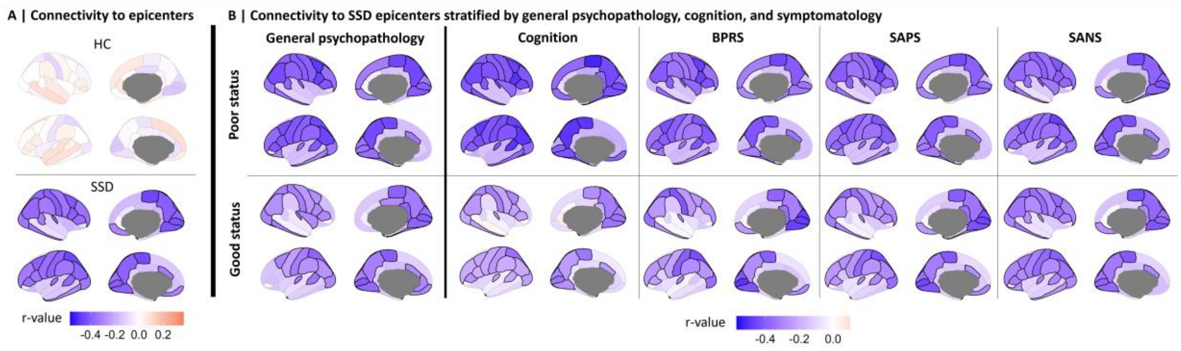

**Supplementary Fig. 12 Connectivity to disease epicenters.** (**A**) Correlation between the corresponding edges to each region and the mean HC (top) and SSD centiles (bottom), where blue and red indicate a negative and positive correlation, respectively. (**B**) Connectivity to SSD epicenters stratified by patients with 'poor' and 'good' general psychopathology, cognition, and symptomatology (BPRS, SAPS, and SANS). The highlighted areas indicate regions where connectivity is significantly correlated with epicenters after tail area-based FDR correction ($P_{Moran} < 0.05$). BPRS, Brief Psychiatric Rating Scale; HC, healthy control; SANS, Scale for the Assessment of Negative Symptoms; SAPS, Scale for the Assessment of Positive Symptoms; SSD, schizophrenia spectrum disorder.

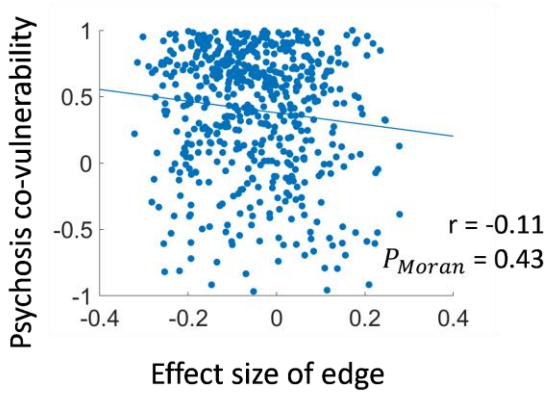

**Supplementary Fig. 13 Correlation (two-sided test) between effect sizes of edges and the structural co-vulnerability to psychosis matrix.**

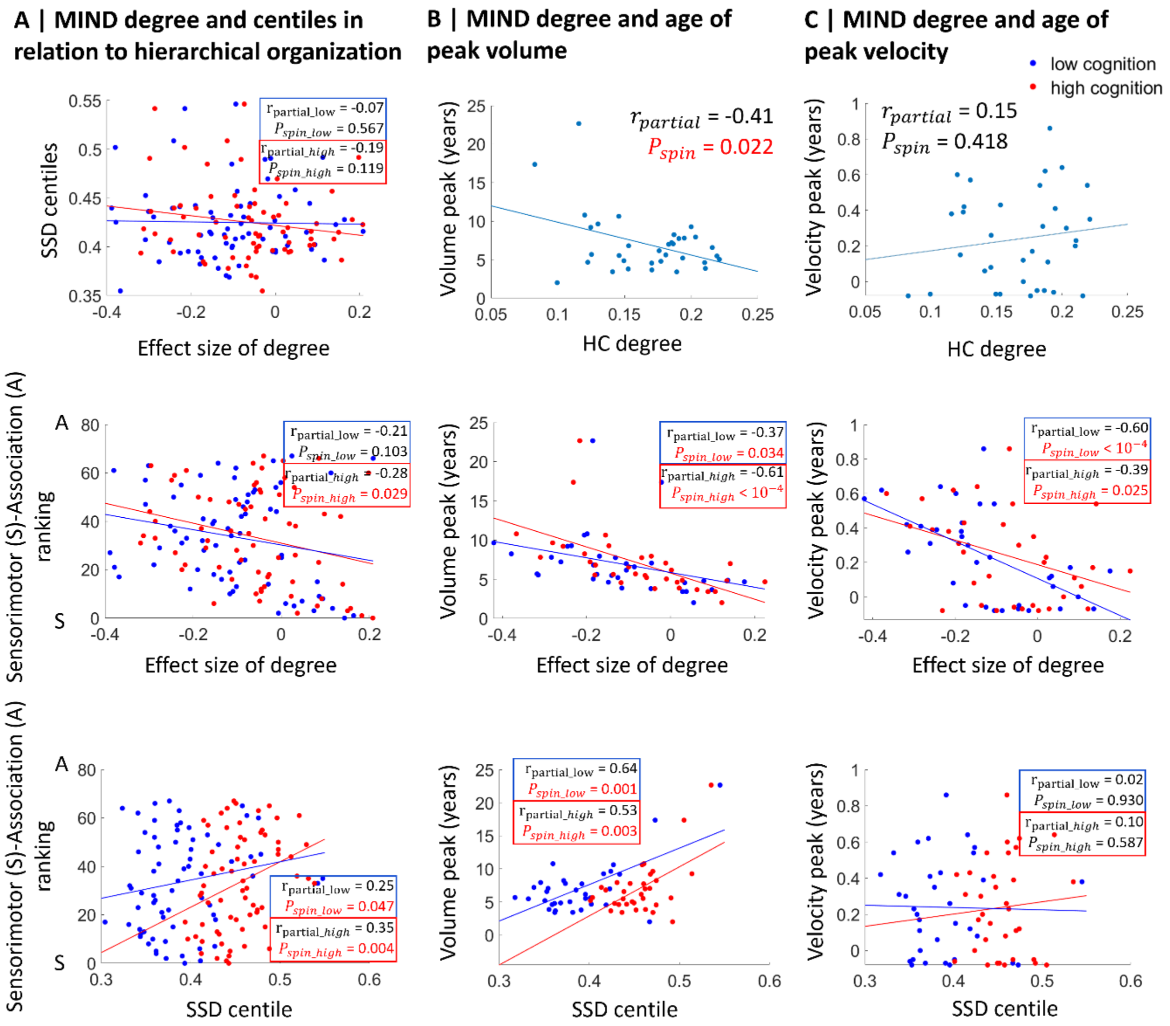

**Supplementary Fig. 14 MIND connectivity stratified by cognition in relation to sensorimotor-association hierarchy and maturational events.** (**A**) Correlations between effect sizes of degrees and SSD centiles (top); effect sizes of degrees and cortical hierarchy (middle); SSD centiles and cortical hierarchy (bottom). (**B**) Correlations between the cortical map of age at regional volume peak (derived from normative trajectories) and HC degree (top); effect size of degree (middle); and SSD centiles (bottom). (**C**) Correlations between the normative cortical map of age at regional velocity peak and HC degree (top); effect size of degree (middle); and SSD centiles (bottom). All regional correlations (two-sided test) accounted for mean regional volumes. HC, healthy control; MIND, Morphometric INverse Divergence; SSD, schizophrenia spectrum disorder.

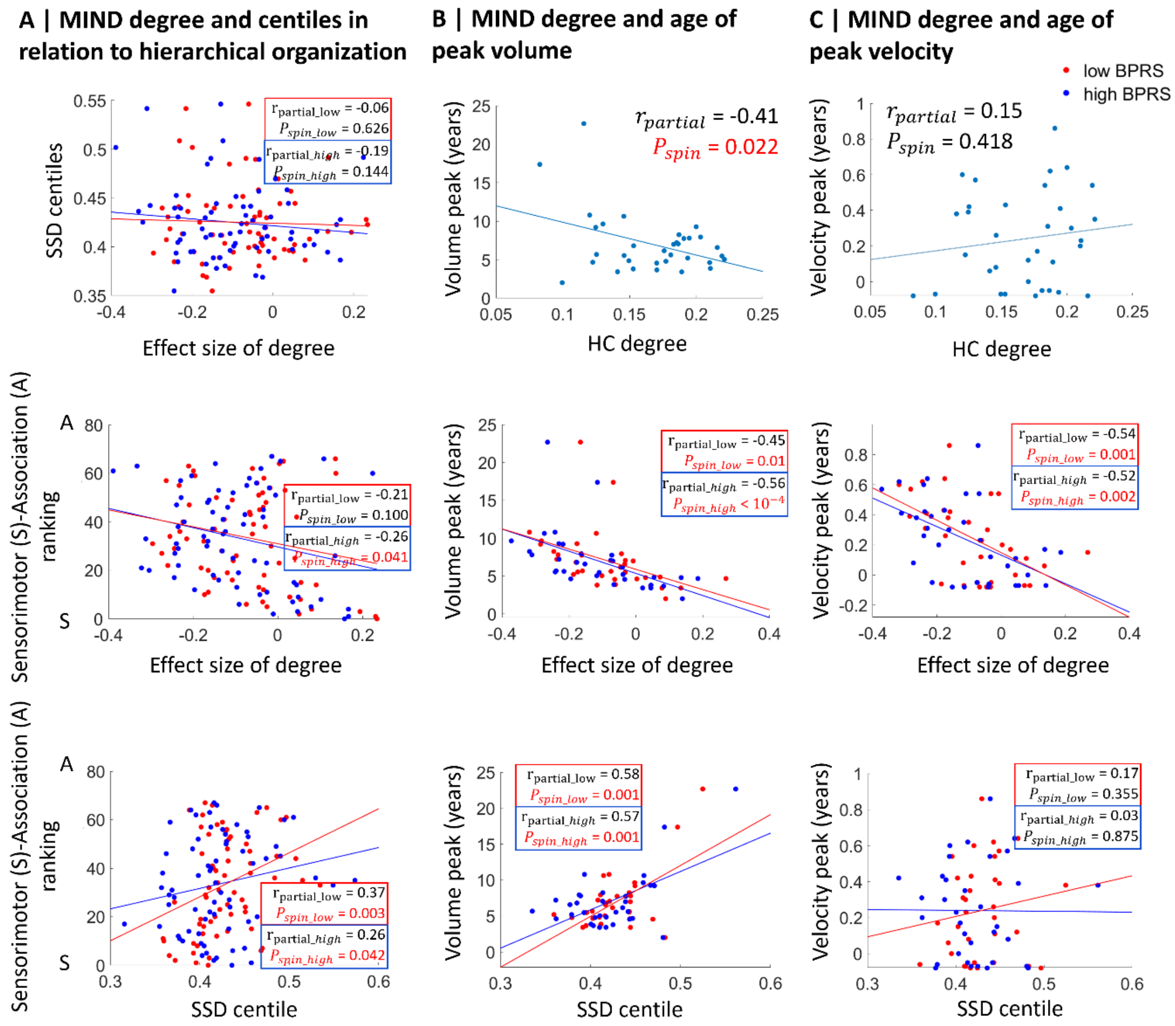

**Supplementary Fig. 15 MIND connectivity stratified by BPRS in relation to sensorimotor-association hierarchy and maturational events.** (**A**) Correlations between effect sizes of degrees and SSD centiles (top); effect sizes of degrees and cortical hierarchy (middle); SSD centiles and cortical hierarchy (bottom). (**B**) Correlations between the cortical map of age at regional volume peak (derived from normative trajectories) and HC degree (top); effect size of degree (middle); and SSD centiles (bottom). (**C**) Correlations between the normative cortical map of age at regional velocity peak and HC degree (top); effect size of degree (middle); and SSD centiles (bottom). All regional correlations (two-sided test) accounted for mean regional volumes. HC, healthy control; MIND, Morphometric INverse Divergence; SSD, schizophrenia spectrum disorder.

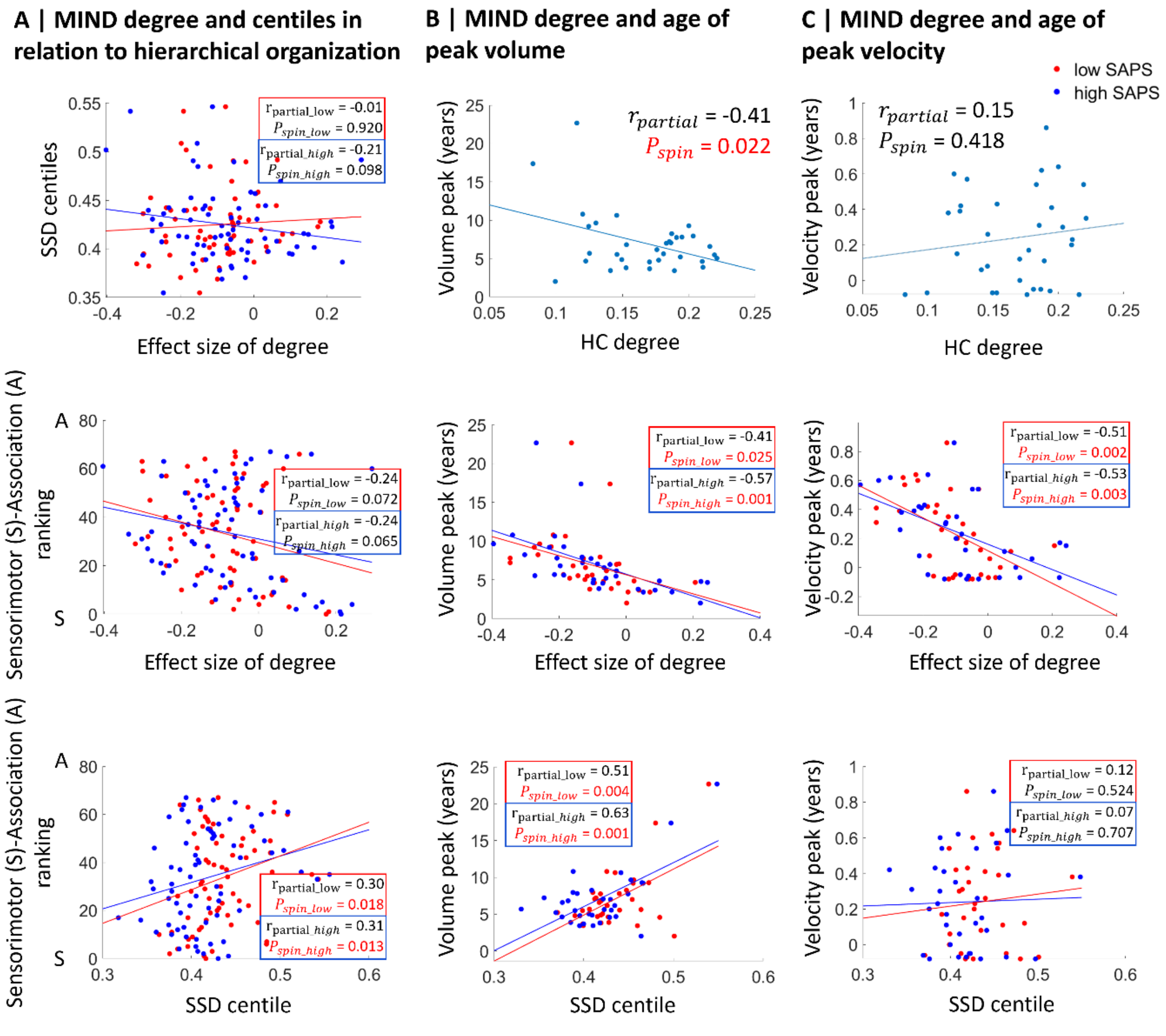

**Supplementary Fig. 16 MIND connectivity stratified by SAPS in relation to sensorimotor-association hierarchy and maturational events.** (**A**) Correlations between effect sizes of degrees and SSD centiles (top); effect sizes of degrees and cortical hierarchy (middle); SSD centiles and cortical hierarchy (bottom). (**B**) Correlations between the cortical map of age at regional volume peak (derived from normative trajectories) and HC degree (top); effect size of degree (middle); and SSD centiles (bottom). (**C**) Correlations between the normative cortical map of age at regional velocity peak and HC degree (top); effect size of degree (middle); and SSD centiles (bottom). All regional correlations (two-sided test) accounted for mean regional volumes. HC, healthy control; MIND, Morphometric INverse Divergence; SSD, schizophrenia spectrum disorder.

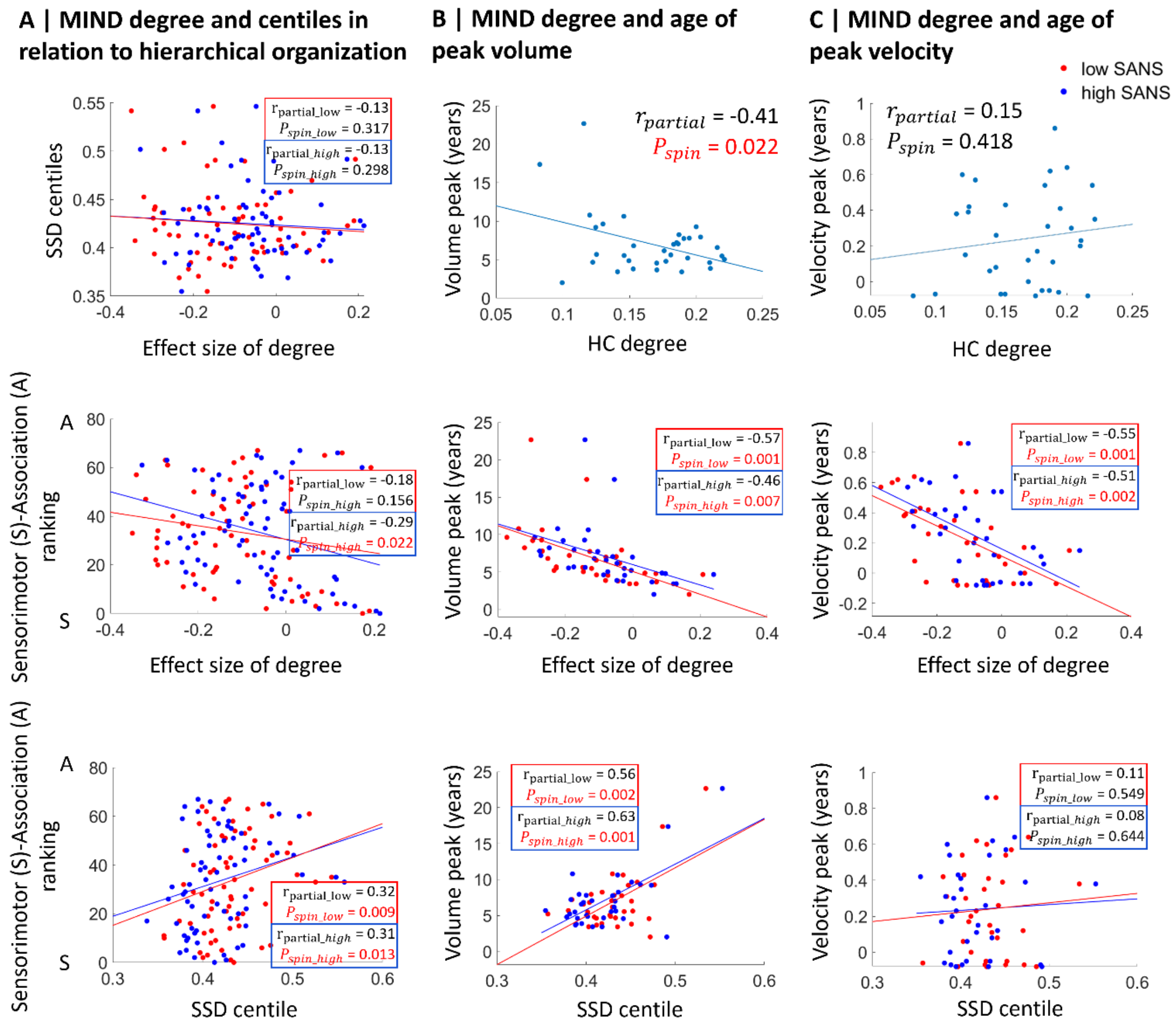

**Supplementary Fig. 17 MIND connectivity stratified by SANS in relation to sensorimotor-association hierarchy and maturational events.** (**A**) Correlations between effect sizes of degrees and SSD centiles (top); effect sizes of degrees and cortical hierarchy (middle); SSD centiles and cortical hierarchy (bottom). (**B**) Correlations between the cortical map of age at regional volume peak (derived from normative trajectories) and HC degree (top); effect size of degree (middle); and SSD centiles (bottom). (**C**) Correlations between the normative cortical map of age at regional velocity peak and HC degree (top); effect size of degree (middle); and SSD centiles (bottom). All regional correlations (two-sided test) accounted for mean regional volumes. HC, healthy control; MIND, Morphometric INverse Divergence; SSD, schizophrenia spectrum disorder.

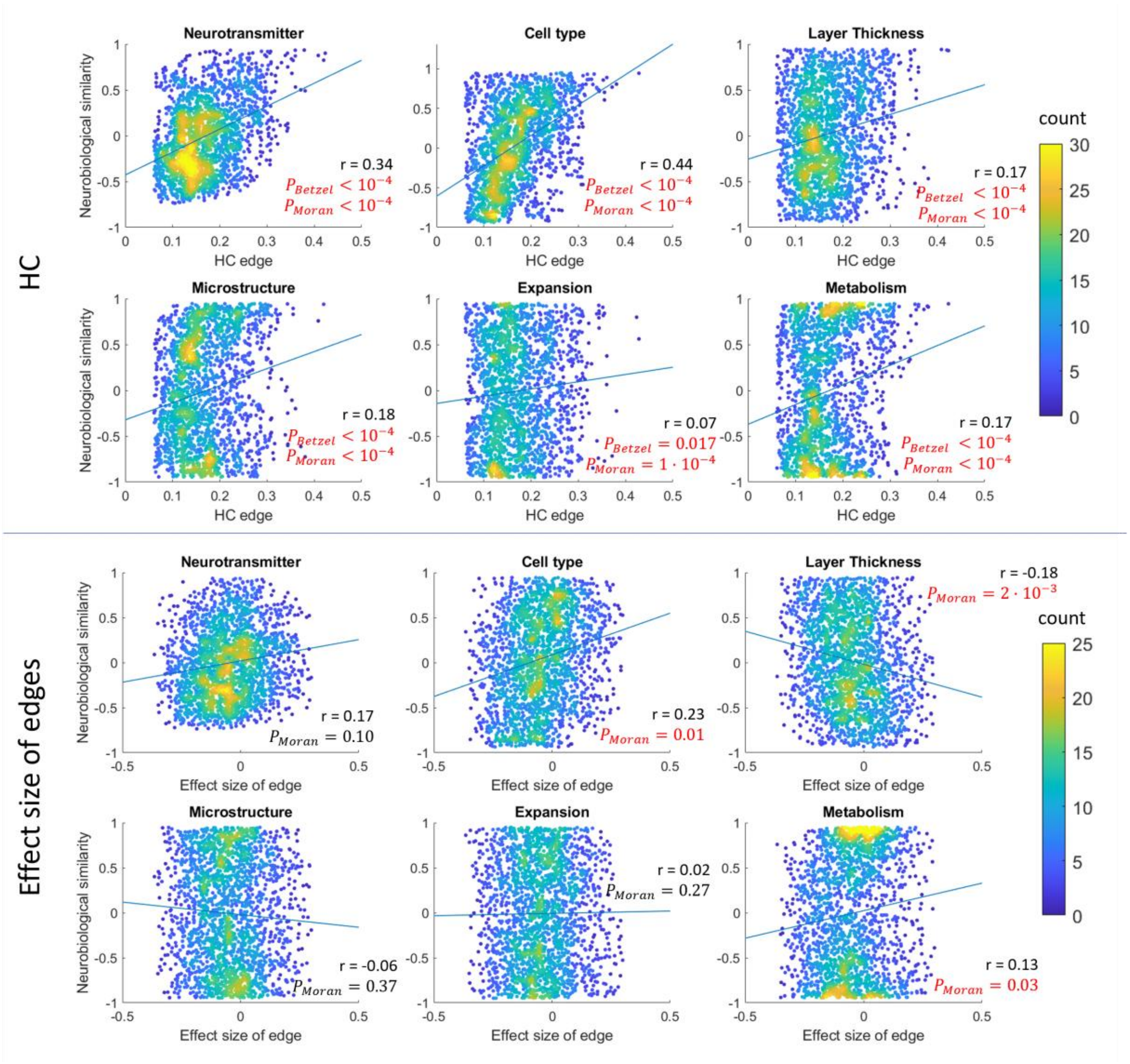

**Supplementary Fig. 18 Neurobiological association of MIND for each neurobiological type.** Association (two-sided test) between the neurobiological similarity matrix and HC edges (top) and effect sizes of MIND edges (bottom). HC, healthy control.

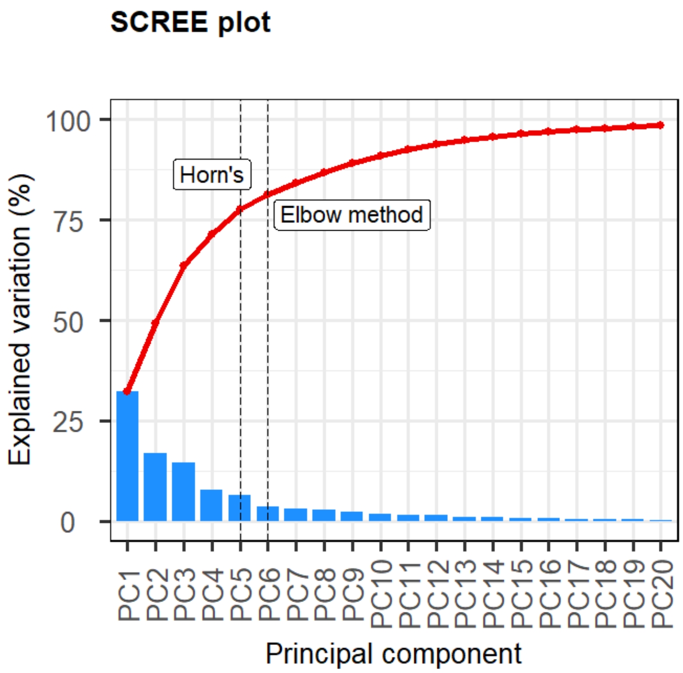

**Supplementary Fig. 19 Cumulative proportion of variance explained by different numbers of principal components derived from the neurobiological matrix.** Horn's parallel analysis and Elbow method were both employed to select the optimum number of PCs to retain in the PCA-CCA analyses.

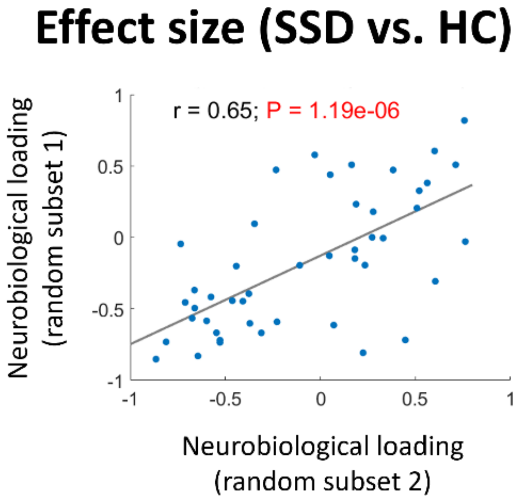

**Supplementary Fig. 20 Correlation between PCA-CCA-derived loadings of each random subset.** HC, healthy control; SSD, schizophrenia spectrum disorder.

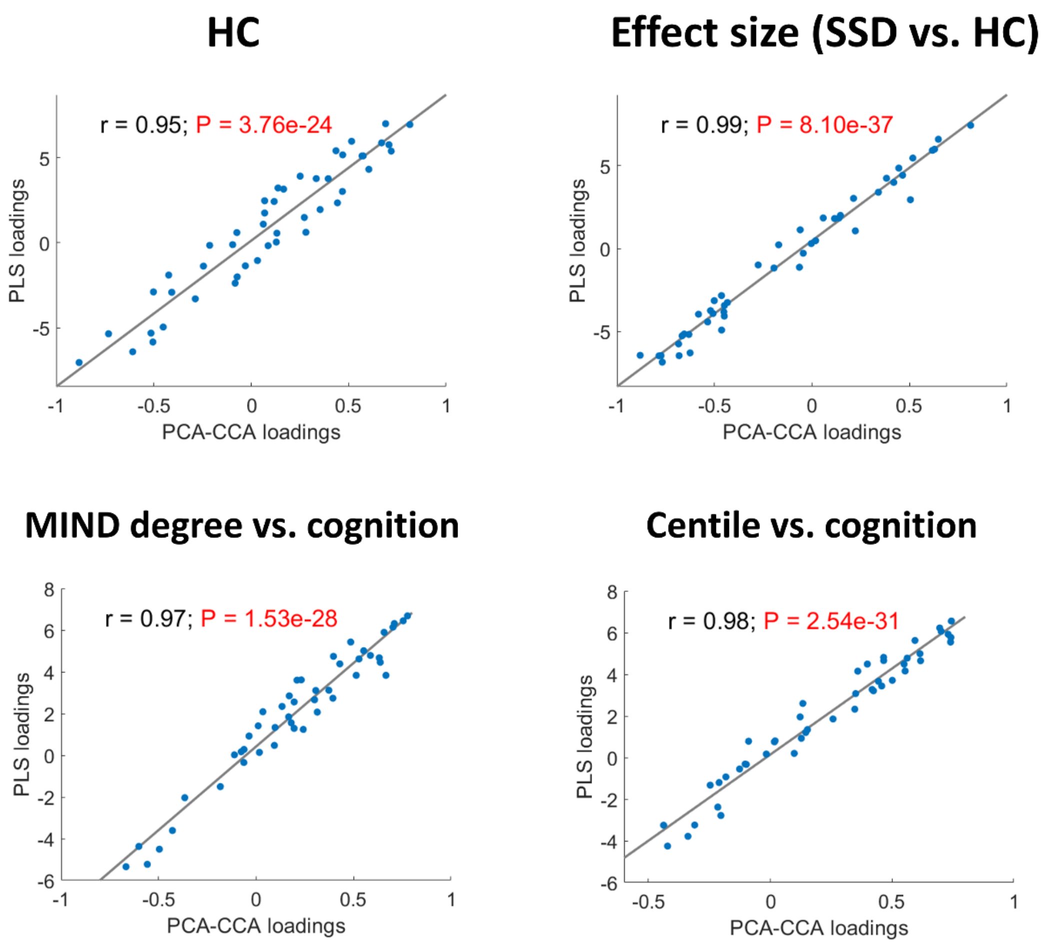

**Supplementary Fig. 21 Correlation between PLS- and PCA-CCA-derived loadings.** HC, healthy control; MIND, Morphometric INverse Divergence; PCA-CCA, Principal Component Analysis-Canonical Correlation Analysis; PLS, Partial Least Square; SSD, schizophrenia spectrum disorder.

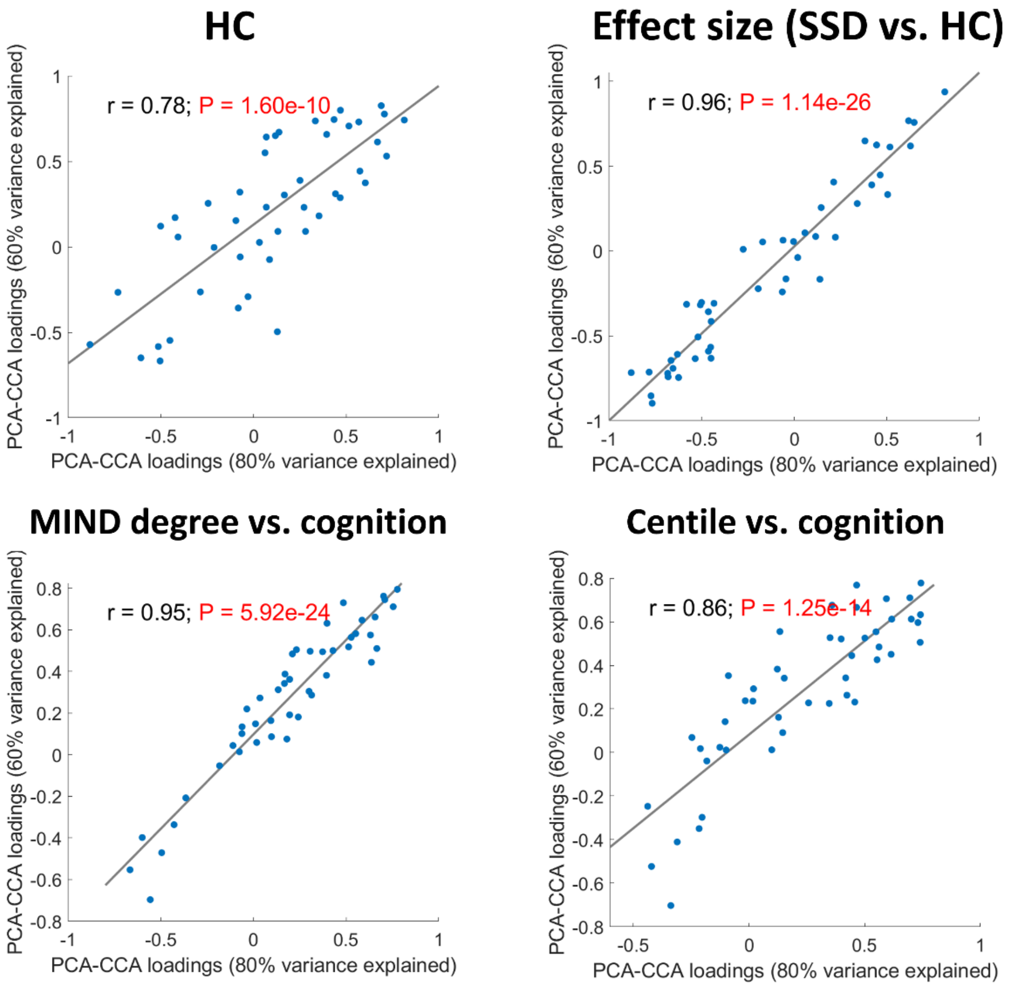

**Supplementary Fig. 22 Correlation between neurobiological loadings derived from PCA-CCA modeling, with explained variances of 60% and 80%.** HC, healthy control; MIND, Morphometric INverse Divergence; PCA-CCA, Principal Component Analysis-Canonical Correlation Analysis; SSD, schizophrenia spectrum disorder.

### Supplemental Tables

#### Supplementary Table 1 Sample description of each psychosis-related group included in the structural co-vulnerability to psychosis matrix

| Group | Diagnosis | n (females) | Age (min, max) | Cohort (n; females) |
|---|---|---|---|---|
| SCZ and SAD-chronic | SCZ | 525 (148) | 37.63 ± 12.06 (10.19, 77.82) | ABCD (2; 1), ASRB (257; 76), BSNIP (100; 30), LA5c (48; 11), MCIC (104; 25), UKB (14; 5) |
| | SAD | 62 (37) | 33.77 ± 10.78 (11.68, 57.06) | B-SNIP |
| FEP | FEP | 352 (138) | 31.43 ± 8.78 (18.78, 59.81) | PAFIP |
| PE | PE-suspected | 48 (33) | 21.55 ± 1.06 (20.78, 24.78) | ALSPAC |
| | PE-definite | 73 (42) | 21.75 ± 1.59 (19.78, 24.78) | ALSPAC |
| | PE-clinical | 36 (28) | 20.89 ± 0.92 (19.78, 23.78) | ALSPAC |
| SCZ and SAD-relatives | SCZ-relatives | 96 (68) | 43.75 ± 15.25 (11.68, 65.07) | B-SNIP |
| | SAD-relatives | 64 (43) | 40.00 ± 16.20 (11.68, 65.07) | B-SNIP |

ABCD, Adolescent Brain and Cognitive Development; ALSPAC, Avon Longitudinal Study of Parents and Children; ASRB, Australian Schizophrenia Research Bank; B-SNIP, Bipolar & Schizophrenia Consortium for Parsing Intermediate Phenotypes; FEP, first-episode psychosis; LA5c, UCLA Consortium for Neuropsychiatric Phenomics LA5c Study; MCIC, Mental Illness and Neuroscience Discovery (MIND) Institute Clinical Imaging Consortium; PAFIP, *Programa de Atención a las Fases Iniciales de Psicosis*; PE, psychotic experience; SAD; schizoaffective disorder; SCZ; schizophrenia; UKB, UK Biobank; .

**Supplementary Table 2 Neurobiological features included in PCA-CCA analyses and the neurobiological similarity matrix**

| Abbreviation | Neurobiological feature | Type |
|---|---|---|
| 5-$HT_{1A}$ | Serotonin receptor | Neurotransmitter |
| 5-$HT_{1B}$ | Serotonin receptor | Neurotransmitter |
| 5-$HT_{2A}$ | Serotonin receptor | Neurotransmitter |
| 5-$HT_4$ | Serotonin receptor | Neurotransmitter |
| 5-$HT_6$ | Serotonin receptor | Neurotransmitter |
| 5-HTT | Serotonin transporter | Neurotransmitter |
| $H_3$ | Histamine receptor | Neurotransmitter |
| $D_1$ | Dopamine receptor | Neurotransmitter |
| $D_2$ | Dopamine receptor | Neurotransmitter |
| DAT | Dopamine transporter | Neurotransmitter |
| NET | Norepinephrine transporter | Neurotransmitter |
| $\alpha_4\beta_2$ | Acetylcholine receptor | Neurotransmitter |
| $M_1$ | Acetylcholine receptor | Neurotransmitter |
| VAChT | Acetylcholine transporter | Neurotransmitter |
| $CB_1$ | Cannabinoid receptor | Neurotransmitter |
| MOR | Opioid receptor | Neurotransmitter |
| $mGluR_5$ | Glutamate receptor | Neurotransmitter |
| NMDA | Glutamate receptor | Neurotransmitter |
| GABA | γ-aminobutyric acid receptor | Neurotransmitter |
| Astro | Astrocytes | Cell type |
| Endo | Endothelial cells | Cell type |
| Micro | Microglia | Cell type |
| Oligo | Oligodendrocytes | Cell type |
| OPC | Oligodendrocyte precursor cells | Cell type |
| Neuro-Ex | Excitatory neurons | Cell type |
| Neuro-In | Inhibitory neurons | Cell type |
| Layer I | Layer I | Layer thickness |
| Layer II | Layer II | Layer thickness |
| Layer III | Layer III | Layer thickness |
| Layer IV | Layer IV | Layer thickness |
| Layer V | Layer V | Layer thickness |

| | | |
|---|---|---|
| Layer VI | Layer VI | Layer thickness |
| Myelin | Myelin | Microstructure |
| Thickness | Cortical thickness | Microstructure |
| Gene PC1 | Gene expression PC1 | Microstructure |
| Neurotransmitter PC1 | Neurotransmitter PC1 | Microstructure |
| Synapse density | Synapse density | Microstructure |
| Evolutionary exp. | Evolutionary expansion | Cortical expansion |
| Developmental exp. | Developmental expansion | Cortical expansion |
| Scaling PNC | Allometric scaling from Philadelphia Neurodevelopmental Cohort | Cortical expansion |
| Scaling NIH | Allometric scaling from National Institutes of Health | Cortical expansion |
| CBF | Cerebral blood flow | Metabolism |
| CBV | Cerebral blood volume | Metabolism |
| $CMRO_2$ | Cerebral metabolic rate of oxygen | Metabolism |
| CMRGlu | Cerebral metabolic rate of glucose | Metabolism |
| Glycolytic index | Glycolytic index | Metabolism |
